# More Than Words: An Integrative Review of Innovative Elicitation Techniques for Qualitative Interviews

**DOI:** 10.1101/2024.05.29.24308062

**Authors:** Renate Kahlke, Lauren A. Maggio, Mark Lee, Sayra Cristancho, Kori LaDonna, Zahra Abdallah, Aakashdeep Khehra, Kushal Kshatri, Tanya Horsley, Lara Varpio

## Abstract

**Introduction:** Interviews are central to many qualitative studies in health professions education (HPE). However, researchers often rely only on oral questioning despite the existence of techniques tailored to elicit the rich data needed to address complex problems and meaningfully engage participants. Elicitation techniques are strategies – e.g. participant photography, neighbourhood walks – used to generate rich conversations, but guidance on these techniques is scattered across literatures from diverse fields. In this synthesis, we offer an overview of the elicitation techniques available and advice about how to choose between them.

**Methods:** We conducted an integrative review, drawing on methodological literature from across the health and social sciences. Our interdisciplinary searches yielded 3056 citations. We included 293 citations that were methodologically focused and discussed elicitation techniques used in interviews with adults. We then extracted specific elicitation techniques, summarizing each technique to capture key features, as well as strengths and weaknesses. From this, we developed a framework to help researchers identify challenges in their interview-based research, and to select elicitation techniques that address their challenges.

**Results:** Elicitation techniques serve two main purposes: they can enrich data and engage participants in new ways. To enrich data, researchers might seek to shift conversations away from participants’ entrenched narratives, to externalize conversations on sensitive topics, or to elicit affect, tacit knowledge, or contextual details. When engaging participants in new ways, researchers might seek to increase equity between the researcher and participant or interview accessibility across diverse participant populations.

**Discussion:** When chosen with study goals in mind, elicitation techniques can enrich interview data. To harness this potential, we need to re-conceptualize interviews as co-production of knowledge by researcher(s) and participant(s). To make interviews more accessible, we need to consider flexibility so that each participant can engage in ways that best suit their needs and preferences.

## Introduction

Qualitative research interviews are mainstay tools for many health professions education (HPE) researchers, and for good reason – if we want to know about our participants’ experiences, it seems intuitive that we should ask them to describe those experiences. Many rich data sets have been generated in this way. However, generating robust interview data is a challenging task. The insights and experiences that researchers seek to explore may be hard for participants to recall or articulate. For example, a topic may be difficult to talk about if it is very sensitive or emotionally loaded for participants. Details may go unstated if participants think they are taken-for-granted, assumed, or trivial, even though they may be key to unlocking deep or novel insights.^1^ Thus, simply prompting people to discuss a topic may not be enough to capture the nuances of their thoughts and experiences. Furthermore, data richness may be limited if oral interviews are not inclusive to all potential participants. For example, participants’ may be challenged to participate in oral interviews if their accent or first language are non-dominant,^2^ if they have cognitive differences that make participation in oral interviews difficult,^3,4^ of if they it difficult to sit, listen, and respond for long periods of time.^5^ Because of long histories of discrimination and disempowerment at the hands of researchers, many historically marginalized potential participants may justifiably decide to not volunteer for interviews, which means that their unique and important perspectives are not part of the research findings informing our HPE practices. Thus, for many studies in HPE, interview data produced from oral interviews may be lopsided— glossing over important details and/or systematically excluding particular groups of participants.

These issues are common, and present challenges for researchers who seek to develop rich data that capture the depth and breadth of experience. But how can researchers tackle these challenges? In many cases, interviewing remains the most appropriate and feasible way to produce data that speak to our research questions; therefore, we suggest that a more robust set of interview techniques is needed. Luckily, there are a wealth of innovative interview elicitation techniques available to researchers. Elicitation techniques are interview approaches that draw on visual, verbal, physical, or written stimuli to help research participants recall specific events and/or articulate their ideas.^1^ These techniques move interviews away from a strict reliance on oral questions and answers, and may include prompts such as photos, concept maps, drawings, and the built environment to generate rich responses to researchers’ queries. These stimuli can be researcher-generated (e.g. participants sort images provided by a researcher and discuss their choices in an interview^6,7^) or – more commonly – participant generated (e.g. participants create drawings^8^ or take photographs^9^ for discussion^10^). In some cases, stimuli are co-constructed by the researcher and participant.^10^ Elicitation interview strategies can be categorized by task—i.e., some techniques rely on constructing (participants discuss something they created), others on arranging (participants arrange and discuss concepts), and yet others on explaining (participants explain their understanding of an object or other stimulus).^1^ Elicitation methods have been lauded for supporting rapport building, facilitating communication, facilitating expression of tacit knowledge, and promoting reflection.^11^

In HPE, researchers have occasionally engaged elicitation techniques to enhance their interviews.^12^ For example, complexity in professional practice has proven difficult to capture using traditional interview methods. To bridge this gap, Cristancho et al.^8^ used rich pictures—a technique where participants draw and then discuss a picture of an event or experience—in interviews with surgeons about complexity in their practice. Similarly, Ajjawi et al.^13^ expanded the field’s thinking about complexity by borrowing video-reflexive interviews from psychology and education, asking participants to reflect on complexity by watching videos of themselves in practice. Other recent examples from HPE include: Dubé et al.’s^14^ guided walk interviews capturing medical students situated experiences of clerkship, Kahlke and Eva’s^15^ researcher-generated concept maps exploring health professions educators’ disparate conceptions of critical thinking, and LaDonna et al.’s^16^ participant-generated photographs illuminating tensions between professional and patient conceptions of health advocacy.

While interview elicitation techniques are beginning to gain traction in HPE research, the vast array of techniques available can be daunting to parse; even the most experienced qualitative researchers may struggle to identify relevant techniques, let alone determine which techniques best suit their topic, methodology, and unique research affordances. To support HPE researchers in enriching their interview data and meeting the challenges of our most complex qualitative research problems, we undertook an integrative review to capture and synthesize interview elicitation techniques used across the health and social sciences. This review asks two questions: 1) What elicitation methods are used within the health and social sciences to enrich interviews? 2) How are they used (for what purpose, in what contexts, and with whom)? From this, we developed a framework to help researchers identify specific challenges in their interview-based research projects and to identify elicitation techniques that will surmount these obstacles. In so doing, we hope to support researchers in designing inclusive research that leads to the production of rich and meaningful interview data.

## Methods

We used an integrative review methodology, a knowledge synthesis approach designed to capture and integrate theoretical, methodological, and/or empirical literatures from multiple fields.^17–19^ To achieve our goal of generating a framework for identifying and selecting interview elicitation techniques, we drew on methodological literature from Anthropology, Education, Health Professions Education, Medicine, Psychology, and Sociology.

Our review takes an interpretive qualitative approach^20–22^ and follows the structure from Whittemore and Knafl’s^19^ integrative review phases: 1) problem identification, 2) literature search, 3) data evaluation, 4) data analysis, and 5) presentation. Problem identification is detailed in the preceding introduction; all other phases are discussed below. Our approach responds to the complexity of a review of this nature, drawing on our team’s extensive experience in qualitative research (RK, KL, SC, LM, LV), review methodologies (LM, TH), and information science (LM).

### Literature Search

In consultation with LM, the information scientist on the research team, we designed complex search strategies optimized for discipline-specific databases where elicitation techniques are regularly used (Psychology: PsycINFO; Education: ERIC; Sociology & Anthropology: Sociological Abstracts), as well as Medline to capture citations from health services and health professions education. Our search strategies used a combination of keywords and controlled vocabulary that included terminology related to elicitation techniques (e.g., photo*, image*, object*) with methodologically focused terminology or within methodologically focused publications (e.g., *International Journal of Qualitative Methods*) (refer to Appendix A: Figure 1 for a complete list of terms and Table 1 for an overview of search strategies). To ensure that we captured elicitation techniques not included in our list of terminology, we also searched title, abstract, keywords for ‘elicit*’ combined with methodological terminology or within a methodological journal.

**Figure 1:**
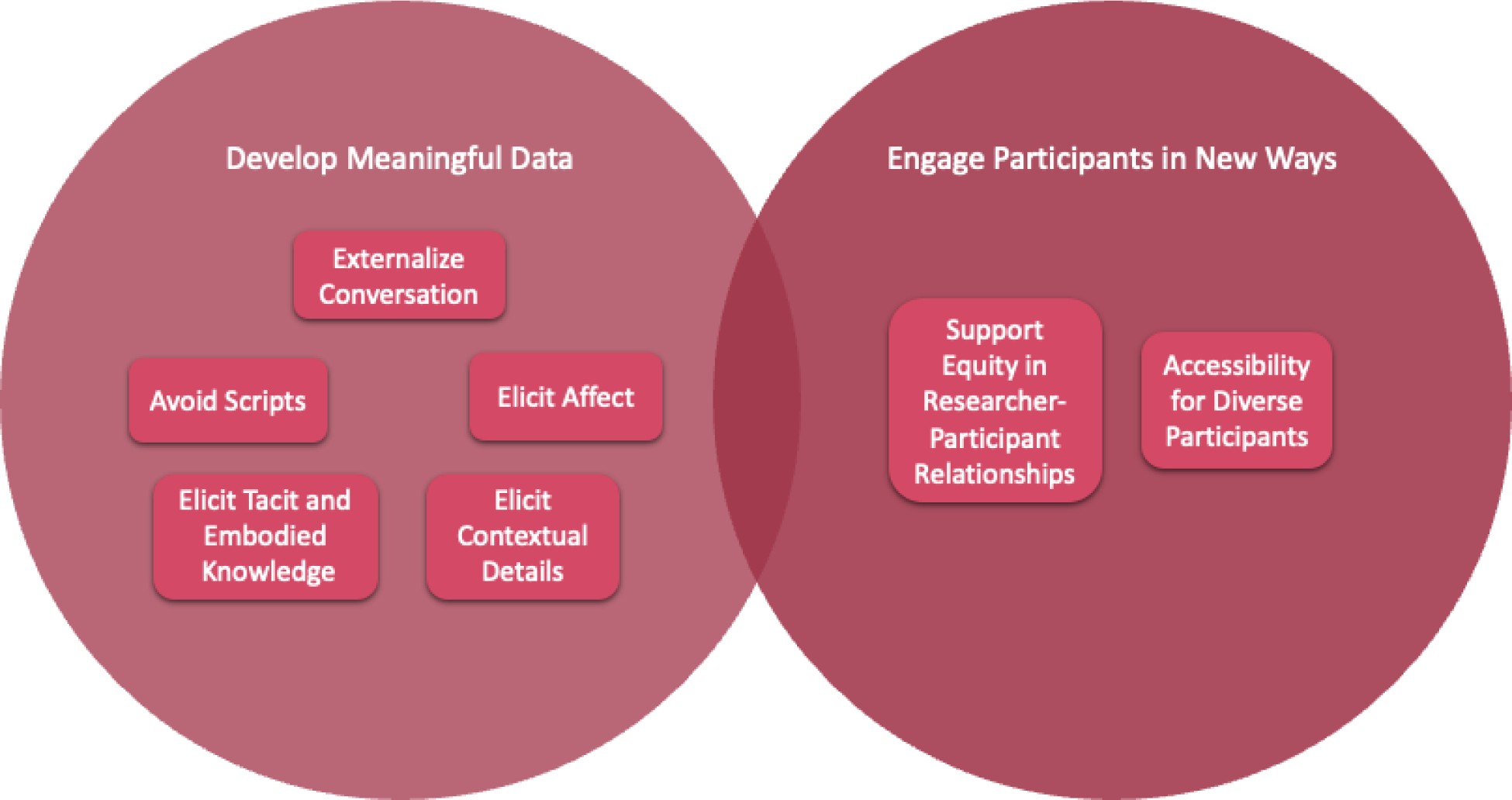
Purposes of Elicitation Techniques. *Note that many techniques can be used in different ways to serve different purposes, or they can be used in one way to serve multiple related purposes.

**Table 1:** Types of Elicitation Techniques.

| Technique | Description | Sub-Techniques | Examples |
| --- | --- | --- | --- |
| Concept mapping | Participants map relationships between concepts/ideas or people | Mind mapping; diagram; network mapping; experience mapping; metaphor; pictor technique; diagrammatic elicitation | Wheeldon J. Is a picture worth a thousand words? Using mind maps to facilitate participant recall in qualitative research<sup>33</sup> |
| Crafting | Participants create tactile art | Collage; fuzzy felt; craft-object making; images; mask-making | Joseph K et al. Unmasking identity dissonance: exploring medial students' professional identity formation through mask making<sup>34</sup> |
| Drawing | Participants draw concepts/experiences | Rich picture; body mapping; postcard; storyboard; cartoon; comic | Molinaro ML et al. Drawing on experience: exploring the pedagogical possibilities of using rich pictures in health professions education<sup>35</sup> |
| Music-elicitation | Participants select, make, or respond to music | Music video | Levell J. 'Those songs were the ones that made me, nobody asked me this question before': music elicitation with ex-gang involved men about their experiences of childhood domestic violence and abuse<sup>36</sup> |
| Object-elicitation | Participants select and/or respond to objects to discuss experiences or values | Object-elicitation; flashcards | Brown N. Identity boxes: Using materials and metaphors to elicit experiences. International Journal of Social Research Methodology<sup>37</sup> |
| Photo and video-elicitation | Participants create photo/video | Photovoice; video-elicitation; video-assisted recall | LaDonna KA et al. Exploring patients' and physicians' perspectives about competent health advocacy<sup>16</sup> |
| Reflective Writing | Participants record reflections/diaries | Reflections; diary writing; solicited diary; solicited audio diary | Monrouxe LV et al. Solicited audio diaries in longitudinal narrative research: A view from inside<sup>38</sup> |
| Sorting | Participants sort items representing concepts | Card Task, repertory grid | Reimer D et al. Pre-clerkship medical students' perceptions of medical professionalism |
| Space and Place Mapping | Participants create or interact with maps (e.g., geographical, architectural) | Mapping | McGrath et al. Building visual worlds: using maps in qualitative psychological research on affect and emotion<sup>39</sup> |
| Storytelling | Participants create stories in different formats | Storyboard, storytelling, cartoon; comic; poetry; drama | Lang et al. Words, camera, music, action: a methodology of digital storytelling in a health care setting<sup>40</sup> |
| Temporal techniques | Participants visually represent experience over time | Timeline drawing; life grid; calendar techniques | Basnet et al. Timeline mapping as a methodological approach to study transitions in health professions education<sup>41</sup> |
| Vignette | Participants respond to researcher-generated vignette | Vignette ; situation-elicitation; story-elicitation; scenario elicitation | Jenkins N, Bloor M, Fischer J, Berney L, Neale J. Putting it in context: the use of vignettes in qualitative interviewing<sup>42</sup> |
| Walking | Participants and interviewer move through places/spaces during interview | Place-elicitation | Dubé TV et al. Interviewing in situ: employing the guided walk as a dynamic form of qualitative inquiry<sup>14</sup> |

Our search process was iterative; as we identified new elicitation techniques in our search results, we refined the search strategies and updated the searches, until the team determined that new searches did not uncover new elicitation approaches. We included peer-reviewed, English-language citations published between April 2010 and 2020. Based on initial exploratory searches, 10 years appeared to capture the bulk of relevant citations. We hand searched reference lists to capture seminal work published prior to this period, as well as any citations specific to the HPE context.^23^ The searches yielded a total of 3056 citations. Refer to Appendix A: Table 2 for complete searches. The original searches were conducted between January and July 2020. In January 2024, we re-searched Medline (2020-2024) – the largest database in our search strategy – to ensure that we had not missed any new approaches to elicitation, yielding 68 new citations. We determined that updating all searches was unnecessary because of the high degree of repetition in techniques within the large number of citations already processed and within the new Medline search, such that our data had sufficient information power to address our research questions.^24^

**Table 2:**
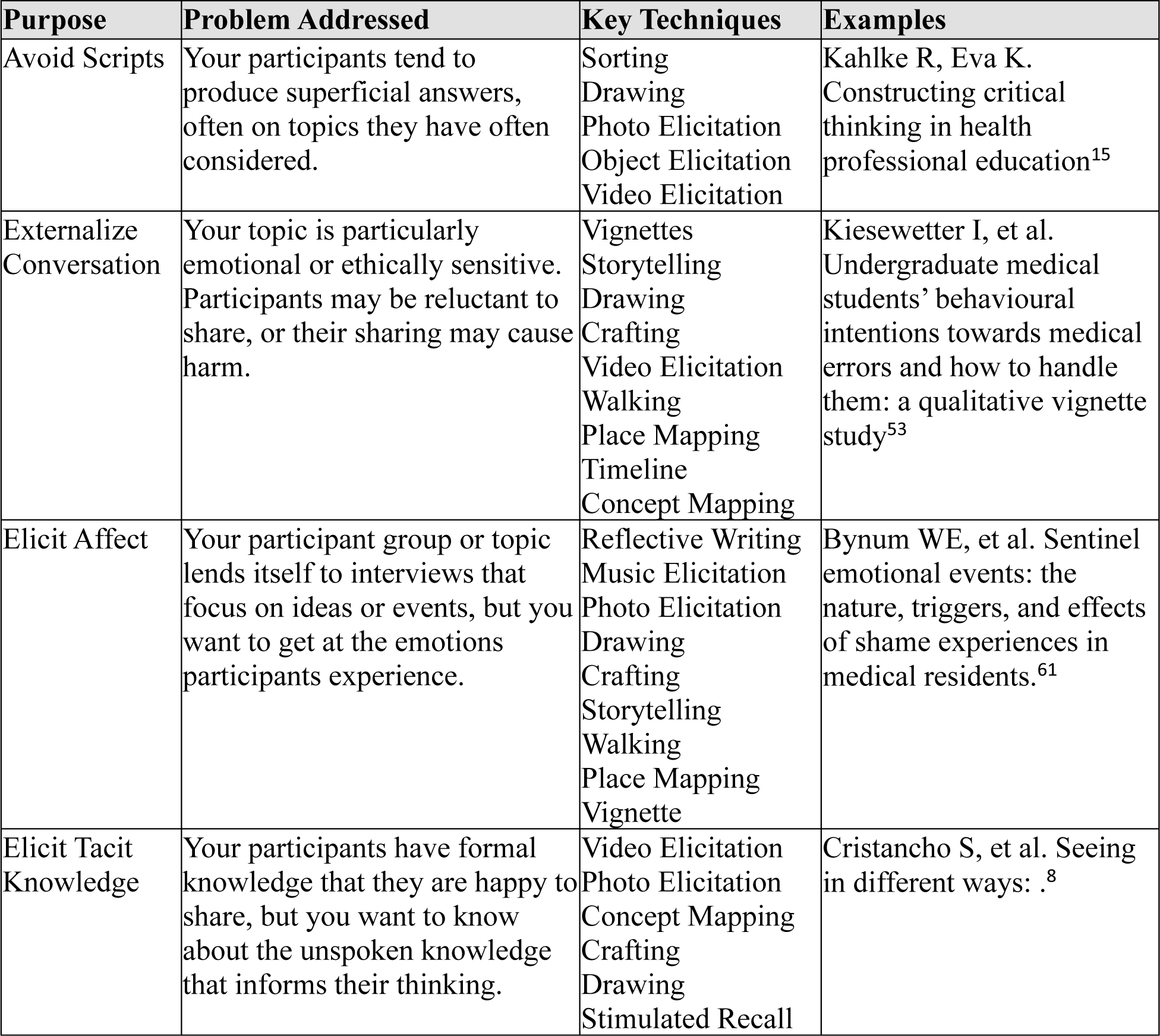
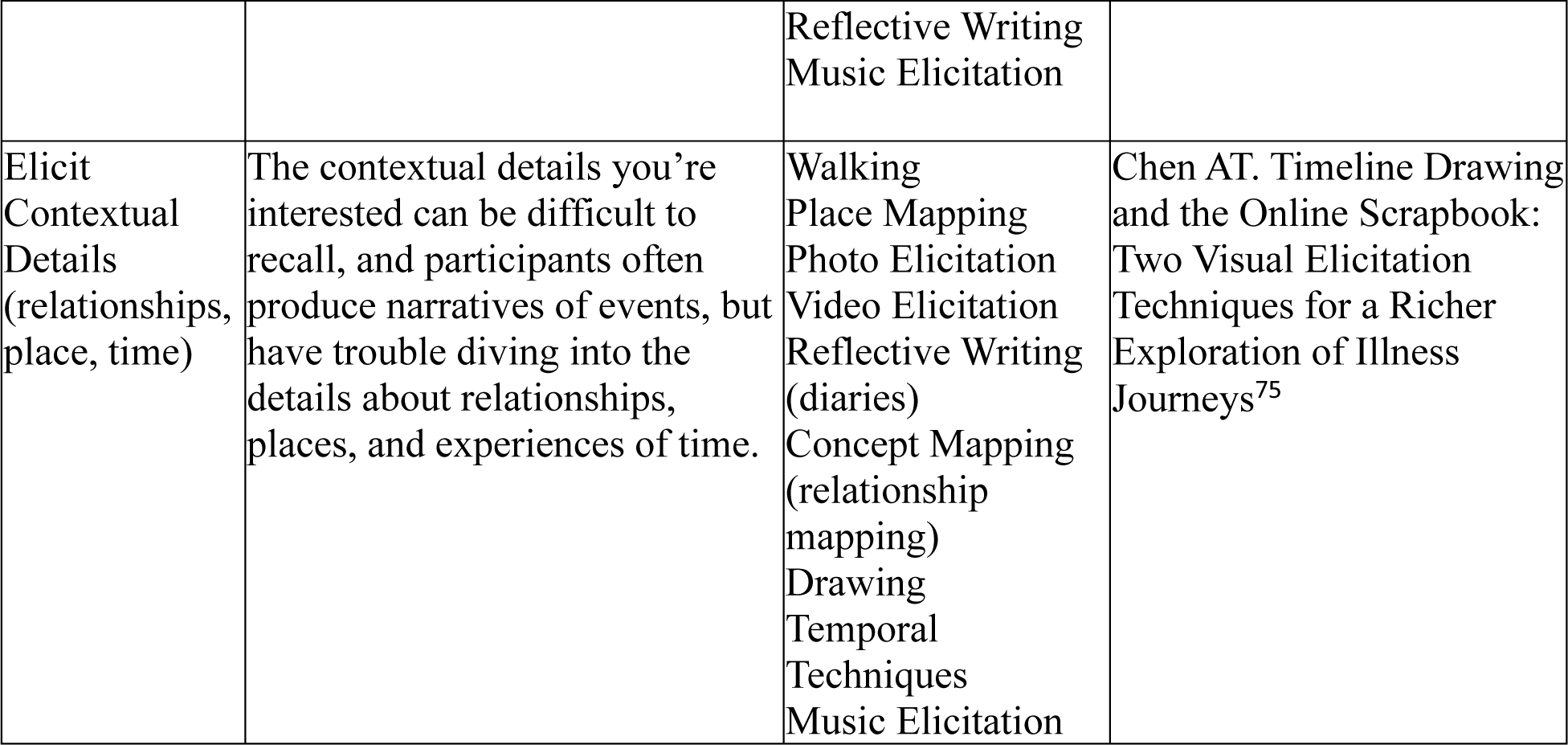
Key Techniques for Developing Meaningful Data. *Although not exhaustive, we have included the most common techniques used for each purpose.

### Data Evaluation

We uploaded the resulting citations to Covidence (an online tool to support data evaluation) for review and duplicates were removed. We then reviewed titles and abstracts and included only records that (a) spoke to a qualitative interview elicitation method and (b) focused on *methodological explication* (i.e., those that foregrounded information about research methods, rather than study results). To be considered eligible for inclusion, we defined interview elicitation methods as any technique, outside of traditional open-ended questions, used in an interview to prompt or structure participant responses (e.g., physical objects, written material, photographs). We excluded articles focused on research with children and youth, as we found that these tended to focus on cognitive and ethical issues involved in working with children, who are not often the focus of HPE research. We also excluded citations when full text was unavailable through our institutions. Article full text was used to determine final inclusion and was assessed independently by at least two reviewers. We met regularly to resolve conflicts and to adjust the search strategy where necessary. Refer to Appendix B for an overview of the data evaluation process.

### Data Analysis

After completing eligibility evaluation, the Principal Investigator (RK) and Research Assistants (RAs) extracted terminology used to describe elicitation techniques (refer to Table 1 below). Members of the research team (RK, KL, SC, LM, LV) then coded each article in the corpus to identify: 1) the role of the participant (i.e., participant-led, researcher-led, or co-constructed); 2) participants’ characteristics (e.g., individuals with communication or cognitive differences); 3) the study design (e.g., epistemology, theoretical perspective, methodology, methods^25^); 4) the type of task (i.e., arrangement, construction, and explanation^1^); 5) the authors’ home discipline(s) and/or field of study. This analysis gave us a broad sense of how, when, where, and why different techniques were used.

Each member of the research team was then assigned two or more sets of techniques for coding and synthesis. Related techniques were combined under a single umbrella if the technique, participant role, participant characteristics, and context were similar. For example, collage-making techniques^26–28^ were combined with other crafting techniques, such as scrapbooking^29^ or feltwork^30^ under the single umbrella of *crafting* elicitation. These techniques are underpinned by similar constructivist epistemologies, with purposes related to engaging with tactile aspects of experience.^29^ Refer to Table 1 for details on each category. In cases where it was difficult to categorize techniques, the PI and RAs discussed, involving other team members where necessary, until we reached agreement.

Articles within each technique were coded by the assigned team member and summarized to capture: (a) the characteristics of each technique, (b) purposes for which it is used, (c) strategies for integrating elicitation prompts using the technique, and (d) strengths and weaknesses identified in the literature. In cases where there were many publications available, the team member sampled from the dataset until they felt they had reached a fulsome understanding of the technique and were no longer encountering new ideas related to the coding categories above. At this point, team members also sought out additional examples from the HPE literature, from our own knowledge and databases, to ensure that we could make results from the broader literature relevant to our HPE research audience.

In keeping with Whittemore and Knalf’s^19^ recommendations, the PI and an RA, in consultation with the senior author (LV), reviewed the summaries and developed a graphic representation of the different uses of elicitation to answer research question 2 (Figure 1).

### Presentation

We used a combination of the SRQR qualitative research reporting guideline^31^ and STORIES reporting guideline for evidence synthesis in healthcare education^32^ to ensure transparent reporting of both the overall review methodology and qualitative analysis.

### Reflexivity

Our team drew on our diverse backgrounds to support our methodology and findings, including information science (LM) and review methodologies (TH, LM). Most of us are also qualitative researchers with an interest in methodological innovation who aim to use these findings to enhance our research and teaching practice (RK, LV, KL, LM), and three are aspiring researchers interested in learning about qualitative research (ZA, AK, KK). Most of us take a social constructionist or subjectivist orientation to our research, which influenced how we approached our questions, the qualitative review methods we used, and our interpretation of results. The fact that we are all knowledge-users is a strength of our team; however, we also acknowledge that our economic and social status as academics (RK, LM, ML, SC, KL, TH, LV) and future health professionals (ZA, AK, KK) may also be a limitation in interpreting and using elicitation techniques with vulnerable populations. We are also all HPE researchers, and while many of us have focused our research on supporting equity-deserving learners and professionals, we primarily engage with the relatively privileged populations of learners and clinicians within the health professions.

## Results

We present our findings in two sections, addressing each of our research questions. First, we offer an overview of techniques we encountered, followed by a framework that delineates the different uses of elicitation techniques in effort to guide researchers in selecting a technique to suit their needs.

### A taxonomy of elicitation techniques

A wide range of elicitation techniques are used to enrich interviews in both the health and social sciences. We constructed a typology of 13 categories of interview elicitation techniques. Table 1 offers a definition for each category, a list of associated terms, and an example publication illustrating a study where the technique was employed. Refer to Appendix C for sub-techniques within each category, and citations for each; Appendix D offers how-to guides for using techniques, where available.

### The purposes of elicitation techniques

We found that interview elicitation techniques can be used to address two broad types of challenges (refer to Figure 1): to develop meaningful data and to engage participants in new ways. Though some techniques have a clear primary aim, most techniques can be used in a variety of ways and serve different purposes (e.g., to elicit embodied knowledge AND to increase accessibility for diverse participants). In Tables 2 and 3, we list the purposes served by interview elicitation techniques; for each purpose we explain the problem these techniques address, record the techniques frequently used under that category, and offer an illustrative example from the published literature.

**Table 3:**
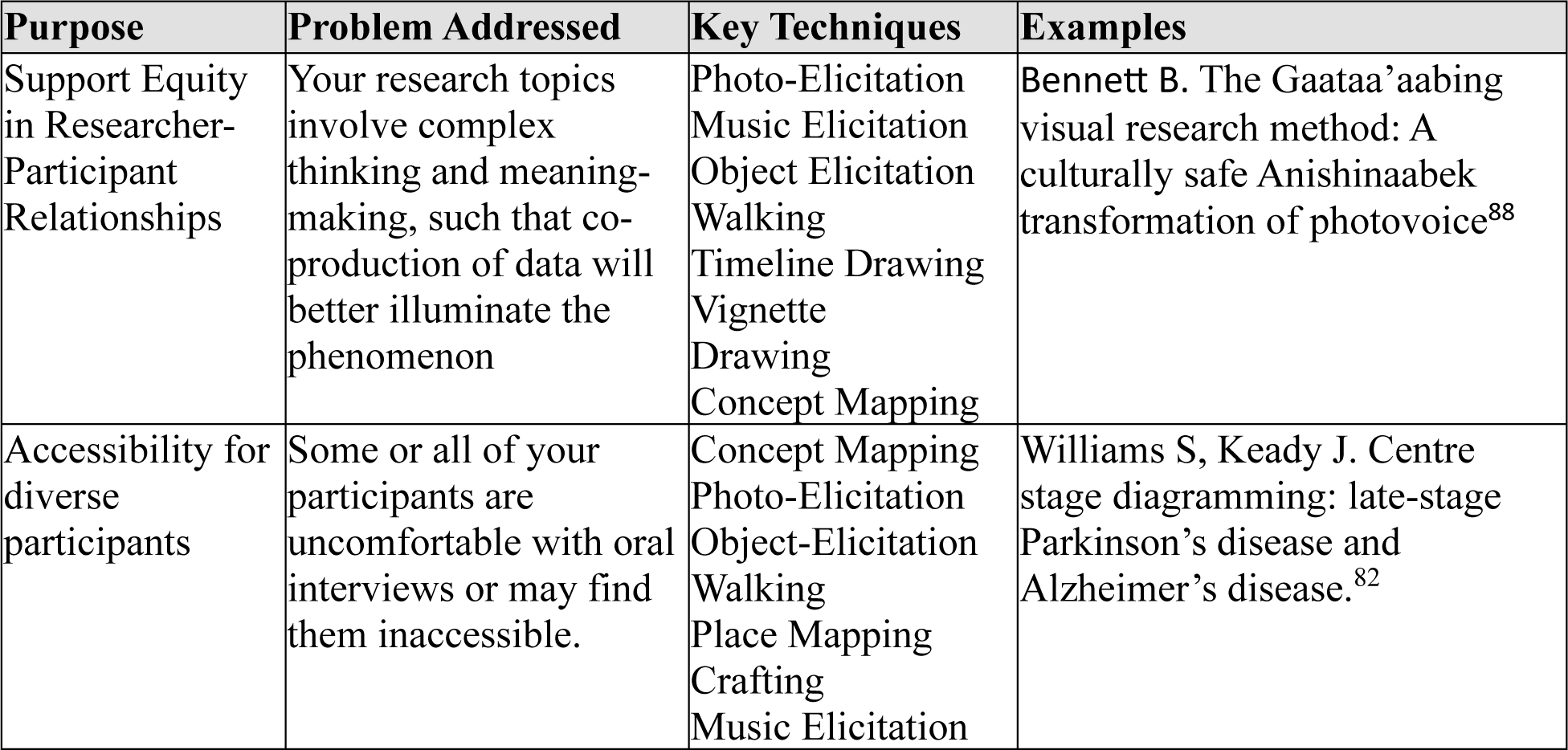
Key Techniques for Engaging Participants in New Ways.

| **Purpose** | **Problem Addressed** | **Key Techniques** | **Examples** |
| --- | --- | --- | --- |
| Support Equity in Researcher-Participant Relationships | Your research topics involve complex thinking and meaning-making, such that co-production of data will better illuminate the phenomenon | Photo-Elicitation<br>Music Elicitation<br>Object Elicitation<br>Walking<br>Timeline Drawing<br>Vignette<br>Drawing<br>Concept Mapping | Bennett B. The Gaataa’aabing visual research method: A culturally safe Anishinaabek transformation of photovoice<sup>88</sup> |
| Accessibility for diverse participants | Some or all of your participants are uncomfortable with oral interviews or may find them inaccessible. | Concept Mapping<br>Photo-Elicitation<br>Object-Elicitation<br>Walking<br>Place Mapping<br>Crafting<br>Music Elicitation | Williams S, Keady J. Centre stage diagramming: late-stage Parkinson’s disease and Alzheimer’s disease.<sup>82</sup> |

### Develop Meaningful Data

First, elicitation techniques are most often described as means for developing meaningful data on topics that are difficult for participants to recall or explain in depth.^1^ More specifically, researchers have used these techniques to: shift participants away from their usual narrations or scripts on a subject, externalize deeply personal or sensitive conversations, elicit affect, discuss tacit or embodied knowledge, or probe for contextual details.

#### Avoid Scripts

Elicitation technique users reported that, for many topics, participants had answers to supply but that these responses can be superficial, not deep reflections on their experiences. By using interview elicitation techniques, researchers were able to shift the interview dynamic, opening new conversations.^1^ For example, object elicitation can focus a conversation on a participant-or researcher-selected object relevant to the topic (e.g. a palliative patient selected a coffee maker to focus a conversation on an aspect of their daily routine important to them^43^); diagrammatic elicitation can be used to invite participants to eschew conventional representations of a phenomenon by creating or responding to diagrams representing a phenomenon (e.g. participants felt better prepared to reflect and identified more unique concepts when they used diagrams to discuss their experiences with a legal reform project^33^).

In HPE, avoiding scripts might be a reason to use interview elicitation techniques when studying phenomena that are common in the field. For example, *critical thinking* is an omnipresent HPE topic and so participants may be prone to repeating existing scripts that reflect common descriptions of critical thinking as a set of generalized skills and attitudes.^44,45^ To avoid collecting normative scripts, one Kahlke and Eva^15^ used object elicitation, asking participants to select an object demonstrating their understanding of *critical thinking* in their teaching. One participant selected a lesson plan, another an artefact representing the history of medicine. Participants and the interviewer then focused the conversation on the object and what it might represent, shifting participants away from their usual conversation patterns on the topic.

#### Externalize Conversation

Researchers often used elicitation techniques to create a focal point for the interview, focusing the conversation on a stimulus, such as participant-selected images^46^ or objects.^43^ This can allow the researcher and participant to focus together and engage in collaborative sense-making, creating new interpretations that neither could produce alone.^43^ When studying particularly emotional topics, elicitation techniques can take pressure off participants and allow them to express ideas without over-focusing on the negative emotional aspects of experience, such as grief or trauma.^42,43,47^ When topics are ethically sensitive, participants can be invited to share their experiences through hypothetical scenarios rather than risking sharing information that could be potentially damaging.^42,46–51^

Vignettes are a key technique used for externalizing conversation because they allow participants to respond in the third person, rather than disclosing personal details.^52^ Vignettes have been used to explore medical students’ thinking about medical errors. This approach created a safe hypothetical space for participants to discuss errors, and avoided the ethical quagmire of handling the potentially unreported errors participants might disclose if asked to focus on their own experience.^53^

#### Elicit Affect

Elicitation techniques can be used to intentionally elicit emotional experiences. These techniques are particularly useful when participants may not be accustomed to talking about or verbally processing emotional aspects of an experience. Rather than recounting events, elicitation techniques can prompt participants to focus on how they felt. For example, participants might be asked to use creative methods to draw places or relationships in which they experienced different emotions,^39,54^ or they might be asked to use photography to capture their emotional experience for discussion.^55,56^

Diaries and other types of reflective writing are often harnessed for capturing emotional experiences.^57–59^ Because diaries are often associated with recording private thoughts and emotions, they can provide an active, reflective space for participants to ‘gather their thoughts.’^60^ For example, Bynum et al.^61,62^ used reflective writing in their research on experiences of shame among medical residents. Participants were invited to write a short reflection on a personal shame experience, which they then discussed with the interviewer. This process offered participants a chance to prepare for an emotionally loaded conversation, and to have agency in the interview process by selecting in advance which experiences they would share.

#### Elicit Tacit Knowledge

Tacit knowledge is comprised of an individual’s meaning making and draws on subjective experience and embodied knowledge.^1^ This type of knowledge can be difficult for participants to narrate in response to direct questioning in an interview; rather, researchers often reported a need to support participants in focussing on their frameworks for thinking about a topic (i.e., tacit knowledge), rather than their formal knowledge. For example, in video-elicitation (also known as stimulated recall), participants can focus on how and why they make decisions in the moment while watching a video recording of a performance in practice. Through this process, participants may notice minute decision-making processes that are not often available retrospectively.^63–65^

In HPE, researchers have used mask-making, an arts-based elicitation technique, to explore professional identity formation among medical students.^34^ Participants were asked to create masks that expressed their changing identity during their medical education; they then explored details of their masks with an interviewer. These researchers were able to capture participants’ knowledge about their identity and identity dissonance that are difficult to render visible through traditional oral interviews. Similarly, researchers have used drawing techniques to explore complexity as encountered by surgeons; by drawing out images of practice complexities, surgeons were able to discuss their practice in new ways, capturing the many intertwining factors that form the tacit knowledge shaping their decisions.^8,66^

#### Elicit Contextual Details

Many elicitation techniques are useful for focusing interviews on details of context that might easily be forgotten, or that participants might not feel are worthy of mention. To capture details about relationships, researchers might invite participants to provide a diagram of their relationships related to a topic (e.g. their support or social networks).^67–70^ Through these diagrams, participants may better describe their relationships and the connections between the people in their lives.^67^ To facilitate conversations about contextual details related to specific places, researchers might ask participants to draw or comment on maps, or to do a walking interview in which interviewer and participant visit the places they discuss.^71–74^ Last, researchers may wish to elicit information related to the temporality of experience through techniques like timeline mapping, where participants map out sequences of events,^75,76^ or diary techniques where participants record their experiences at particular moments in time, often with a focus on longitudinal data generation.^60,77^

In medical education, walking interviews have been used with success to investigate the experiences of medical students, within their living and placement contexts.^14^ This approach allowed researchers to deepen their understanding of medical students’ lived experience of place, probing for clarification on the importance of different aspects of students’ environments.

### Engage Participants in New Ways

A second broad purpose for using interview elicitation techniques is to engage participants in new ways. Researchers might seek to use elicitation techniques to shift the research-participant relationship to empower participants, or they might seek to engage participants otherwise excluded from research processes.

#### Support Equity in Researcher-Participant Relationships

In many oral interviews, researchers have significant control over the topic and flow of the interview, maintaining researcher-participant hierarchies. Many authors in our review note that decentering these hierarchies is a significant benefit involved in using participant-directed elicitation techniques. For example, when participants decide when and what is relevant for their diary entries,^77,78^ select objects for discussion,^43^ or draw maps of their environment highlighting things they see as important,^79–81^ they often feel empowered to focus the interview on experiences that they want to share, rather than solely on the experiences the interviewer deems important.^1^ The process of focusing interview conversations on elicitation stimuli can allow interviewer and participant to co-create shared meanings; when participants are given greater control over the stimuli discussed, these conversations are further equalized to create a foundation for shared meaning-making.

For example, music elicitation can allow participants to control both the tone and topics of interview conversations. Interviewers can invite participants to select – or even create^82^ – music related to the research topic, offering control over what would be discussed and setting the tone for the interview.^36,83^ In HPE, music elicitation could be used to elicit participant accounts of their emotions post-call, inviting participant control over the interview when they select music that expresses the emotional experiences that they wish to discuss.

#### Accessibility for Diverse Participants

Finally, elicitation techniques can be used to engage participants who do not always feel empowered to participate in research. Many techniques have been used to enhance participant control over the research process and encourage participation, particularly for vulnerable groups, such as those who use drugs.^84,85^ For such participants, it is often difficult to trust researchers with data – in the wrong hands such data can lead to professional or legal consequences and increase their vulnerability. There are populations in HPE who are vulnerable—e.g., racialized learners who have experienced mistreatment and fear reprisal when they disclose details about these experiences.^86^ These participants may be rightfully concerned about how their data will be presented and used. For these participants, control over the research process can be key to participation.

Additionally, elicitation techniques can be used to overcome barriers in accessibility. For example, graphic elicitation has been used with patients who have memory- or attention-related challenges. The diagrams participants generate during the interview can help focus attention and ensure that the participant sets the pace of the interview.^87^ In the HPE context, interviews focused on elicitation stimuli, such as photographs or objects, may encourage engagement from patients and learners who live with sensory processing issues that make oral interviews difficult.

## Discussion

While oral interviews are often used to tackle problems relevant to HPE researchers, the rich responses researchers seek are not always easily elicited with this method. Participants may have front-of-mind responses, or scripts, that are readily produced at the expense of deeper insights, and topics may be too sensitive to tackle head-on. Participants do not always find it easy to narrate their affective experience, tacit knowledge, or the rich contextual details surrounding their experiences. Additionally, oral interviews often reproduce power imbalances between interviewer and participants – stifling possibilities for shared meaning-making or preventing some participants from engaging at all. However, as our data demonstrate, different elicitation techniques can powerfully influence interview dynamics.

Many of these techniques require a different way of thinking about data production, one that moves away from preoccupations with consistency and replicability in data collection. Instead, most of the elicitation techniques described in this article embrace a social constructionist stance in which researchers are never neutral and meaning is co-constructed through the interview process.^25,89^ Data are not collected, but co-produced and, as a result, are unique to the situation (e.g. individual interviewer, participant, place, and time). When we empower participants to take more control over the interview process via varied elicitation techniques, consistency is not only unachievable but, more importantly, it is also not desirable.

To enable potential participants’ full engagement, we need to think differently about inclusion in research design. This will require focusing on engaging a broader range of participants and creating more inclusive interview formats that allow participants to express their expansive experiences. For such inclusion, HPE researchers can turn to Universal Design for Research (UDR),^90^ defined as “the design of research so that all people can be included as potential participants, to the greatest extent possible, without the need for adaptation or specialized design.”^91^ Key to engaging with UDR in qualitative interviewing is a focus on multisensory and flexible options for participants, baked into the research design.^91^ This means a person-centred approach to interviewing, in which researchers and participants can choose interview approaches to meet their unique needs, preferences, and situations.^3^ For example, within a single study, some participants may prefer to lead a walking interview that empowers them to control the interview environment, while others may want to involve photographs or other visual materials to address challenges they might have with focus or expression.^3^

For many researchers, including this research team, this variability in the type of data produced can create analytic discomfort. Many of us are only trained to analyze words, and it can be daunting to face the challenge of analyzing diverse participant-directed interview formats as well as artefacts such as images, objects, or music that defy our existing modes of analysis. HPE methodological literature to support this shift are few and far between,^66,92–94^ but resources are available in the literatures of other disciplines. For example, instead of analyzing only our interview transcripts, researchers can engage with the icons and symbols in participant-produced drawings through iconographic analysis, originating in the field of art criticism.^66^ Alternatively, semiotic analysis of videos might involve attending to visual details embedded in gestures and movements.^92^ While such resources exist, HPE researchers who decide to use the elicitation techniques described in this manuscript would be well advised to seek out a consultation or collaboration from scholars who are savvy users of the specific technique selected.

To capitalize on the richness of multiple forms of data, empower diverse participants through UDR, and unlock the potential of co-construction, we argue for letting go of outdated beliefs that consistency is necessary for rigour and that words are more valuable than other forms of meaning-making.^94^ This will not be easy. Not only will it require learning new approaches to interviewing and data analysis, but it will also involve educating editors and reviewers who will likely be unfamiliar with these approaches.

## Conclusions

Novel interview elicitation methods – ranging from researcher-generated vignettes to participant generated collages – can help HPE researchers produce rich and nuanced data from a wide range of research participants. These approaches come with both immense potential to create novel insights and challenges to the way we currently think about qualitative HPE research. To harness the potential of many elicitation techniques we must think differently about how we engage with participants and how we analyze the diverse data we produce.

## Data Availability

All data are available online at

https://doi.org/10.5281/zenodo.11357567

## Acknowledgements

The authors would like to thank the Society of Directors of Research in Medical Education for funding this work, and Lindsey Sikora for her support in crafting and refining our early searches.

## Notes

The authors have no conflicts of interest to disclose.

### Competing Interest Statement

The authors have declared no competing interest.

### Funding Statement

Support was received from the Society of Directors of Research in Medical Education

