## Appendix A for "More Than Words: An Integrative Review of Innovative Elicitation Techniques for Qualitative Interviews"

### Search Strategies

Our team brainstormed an initial list of terminology related to elicitation techniques. Refer to Figure 1 below for complete list, including a timeline for addition of new terms, based on findings from our initial searches. To support our searches, we selected three discipline-specific databases most relevant to interview elicitation (PsychINFO, ERIC, SocAbstracts).

We then employed three search strategies within these three databases: 1) we searched for methodological journals (‘qualitative’ or ‘method*) AND our specific elicitation terms as subject (expert indexed where available); 2) we searched for articles that were expert-indexed using methodological terms (e.g. mainsubject) AND our specific elicitation terms as subject or mainsubject (where available); 3) to capture terms that were not included in our list of specific technique terms, we searched for articles that were expert-indexed using methodological terms as the main subject AND included “elicit*” anywhere in the keywords, title, or abstract (everywhere but full text).

To capture terms that might appear outside of our three disciplinary databases, as well as uses of elicitation techniques in medicine and medical education, we added a fourth strategy: 4) searched Medline using the MESH term “Interviews as Topic” AND our specific technique terms. These 4 strategies are detailed in Table 1. Refer to Table 2 for full search strings related to each search strategy.

**Fig 1: Specific Technique Search Terms and Screening Process**

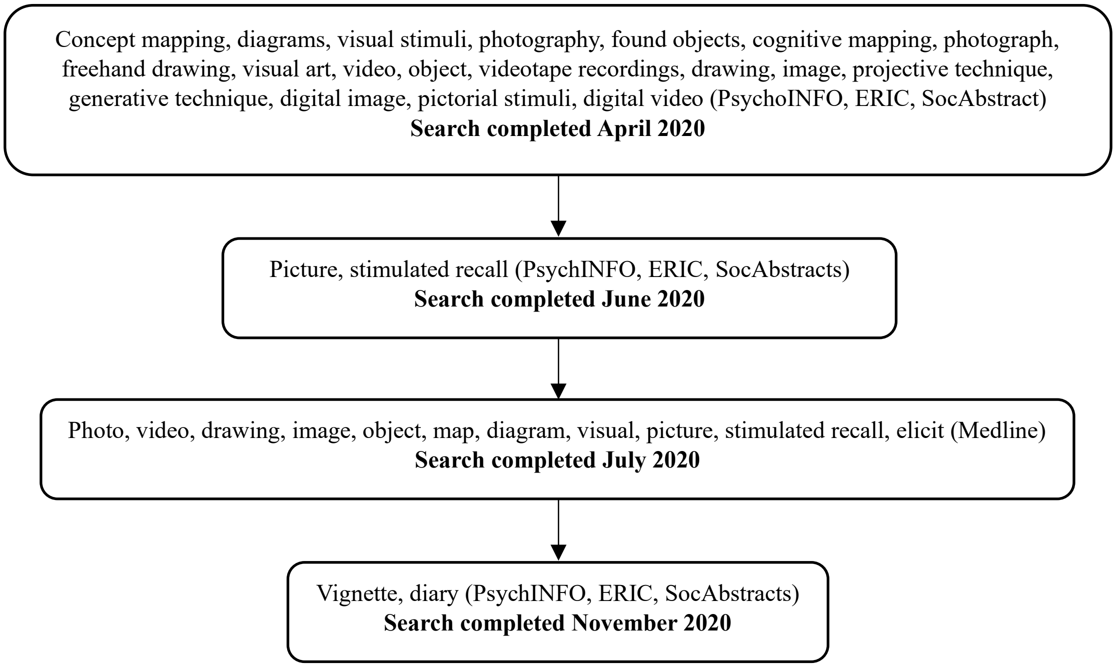

**Table 1: Four search strategies**

| Search Strategy | Databases | Terms To Capture Methodological Publications | | Terms to Capture Elicitation Techniques |
|---|---|---|---|---|
| 1 | PsychINFO, ERIC, SocAbstracts | Journal = ‘qualitative’ OR ‘method*’ | AND | Subject = Specific technique terms |
| 2 | PsychINFO, ERIC, SocAbstracts | Subject = methodological terms (method, data collection, etc) | AND | Subject = Specific technique terms |
| 3 | PsychINFO, ERIC, SocAbstracts | Subject = methodological terms (method, data collection, etc) | AND | Search everywhere except full text: elict* |
| 4 | Medline | MESH = Interviews as Topic | AND | Subject = Specific technique terms |

**Table 2: Search Strings Related to Four Search Strategies**

| Search Strategy | Databases | Example Search Strings |
|---|---|---|
| 1 | PsychINFO | ((visual method* or map* or diagram* or projective technique* or generative technique*).id. or (visual feedback or photographs or digital images or pictorial stimuli or digital video or video-based interventions or videotapes or human figures drawing or projective personality measures or cognitive maps).sh.) and ((qualitative or method*).jn. or (qualitative or method*).bt.) and (abstracts and ("0110 peer-reviewed journal" or "0130 peer-reviewed status unknown" or "0200 book" or "0240 authored book" or "0280 edited book" or "0300 encyclopedia") and yr="2009 -Current") |
| 1 | ERIC | (mainsubject.Exact("concept mapping" OR "diagrams" OR "visual stimuli" OR "photograph" OR "found objects" OR "cognitive mapping" OR "photographs" OR "freehand drawing" OR "visual arts" OR "photography") OR su(video* OR diagram* OR object* OR map* OR visual*)) AND (pub(qualitative OR method*)) |
| 1 | SocAbstracts | (mainsubject.Exact("videotape recordings" OR "videotapes" OR "map/maps/mapped/mapping" OR "videotape recording" OR "video" OR "visual stimuli" OR "drawings" OR "videotaped interviews" OR "drawing" OR "image" OR "images" OR "concept |

| | | |
|---|---|---|
| | | mapping" OR "mapping" OR "video recording" OR "image/images" OR "cognitive maps" OR "video recordings" OR "object/objects" OR "cognitive mapping" OR "photographs" OR "draw/draws/drawing/drawings" OR "photography" OR "photography/photographic" OR "video recorders") OR su(visual* OR diagram*)) AND pub(qualitative OR method*) |
| 2 | PsychINFO | ((visual method* or map* or diagram* or projective technique* or generative technique*).id. or (visual feedback or photographs or digital images or pictorial stimuli or digital video or video-based interventions or videotapes or human figures drawing or projective personality measures or cognitive maps).sh.) and (methodology or qualitative methods or focus groups or semi-structured interview or data collection or interviews or qualitative measures).sh. and (abstracts and ("0110 peer-reviewed journal" or "0130 peer-reviewed status unknown" or "0200 book" or "0240 authored book" or "0280 edited book" or "0300 encyclopedia") and yr="2009 -Current") |
| 2 | ERIC | (mainsubject.Exact("concept mapping" OR "diagrams" OR "visual stimuli" OR "photograph" OR "found objects" OR "cognitive mapping" OR "photographs" OR "freehand drawing" OR "visual arts" OR "photography") OR su(video* OR diagram* OR object* OR map* OR visual*)) AND (mainsubject.Exact("research design" OR "research methodology" OR "data collection" OR "methods research" OR "research tools")) |
| 2 | SocAbstracts | (mainsubject.Exact("videotape recordings" OR "videotapes" OR "map/maps/mapped/mapping" OR "videotape recording" OR "video" OR "visual stimuli" OR "drawings" OR "videotaped interviews" OR "drawing" OR "image" OR "images" OR "concept mapping" OR "mapping" OR "video recording" OR "image/images" OR "cognitive maps" OR "video recordings" OR "object/objects" OR "cognitive mapping" OR "photographs" OR "draw/draws/drawing/drawings" OR "photography" OR "photography/photographic" OR "video recorders") OR su(visual* OR diagram*)) AND mainsubject.Exact("methodology (data collection)" OR "methodology/methodologies/methodological (see also method)" OR "research design" OR "qualitative methods" OR "methodology/methodologies/ methodological (see also method)" OR "focus groups" OR "focus group interviews" OR "interviews as topic" OR "data collection" OR "visual sociology") |
| 3 | PsychINFO | elicit*.tw. and (methodology or qualitative methods or focus groups or semi-structured interview or data collection or interviews or qualitative measures).sh. and (abstracts and ("0110 |

| | | |
|---|---|---|
| | | peer-reviewed journal" or "0130 peer-reviewed status unknown" or "0200 book" or "0240 authored book" or "0280 edited book" or "0300 encyclopedia") and yr="2009 -Current") |
| 3 | ERIC | (noft(elicit*)) AND (mainsubject.Exact("research design" OR "research methodology" OR "data collection" OR "methods research" OR "research tools")) |
| 3 | SocAbstracts | noft(elicit*) AND mainsubject.Exact("methodology (data collection)" OR "methodology/methodologies/methodological (see also method)" OR "cognitive interviews" OR "research design" OR "qualitative methods" OR "methodology/methodologies/ methodological (see also method)" OR "focus groups" OR "focus group interviews" OR "interviews as topic" OR "data collection" OR "visual sociology") AND stype.exact("Books" OR "Scholarly Journals") AND PEER(yes) |
| 4 | Medline | ((photo* or video* or drawing* or image* or object* or map* or diagram* or visual or picture* or stimulated recall or elicit*) and interview).sh. |
