## Appendix B for "More Than Words: An Integrative Review of Innovative Elicitation Techniques for Qualitative Interviews"

### Screening Process using Covidence

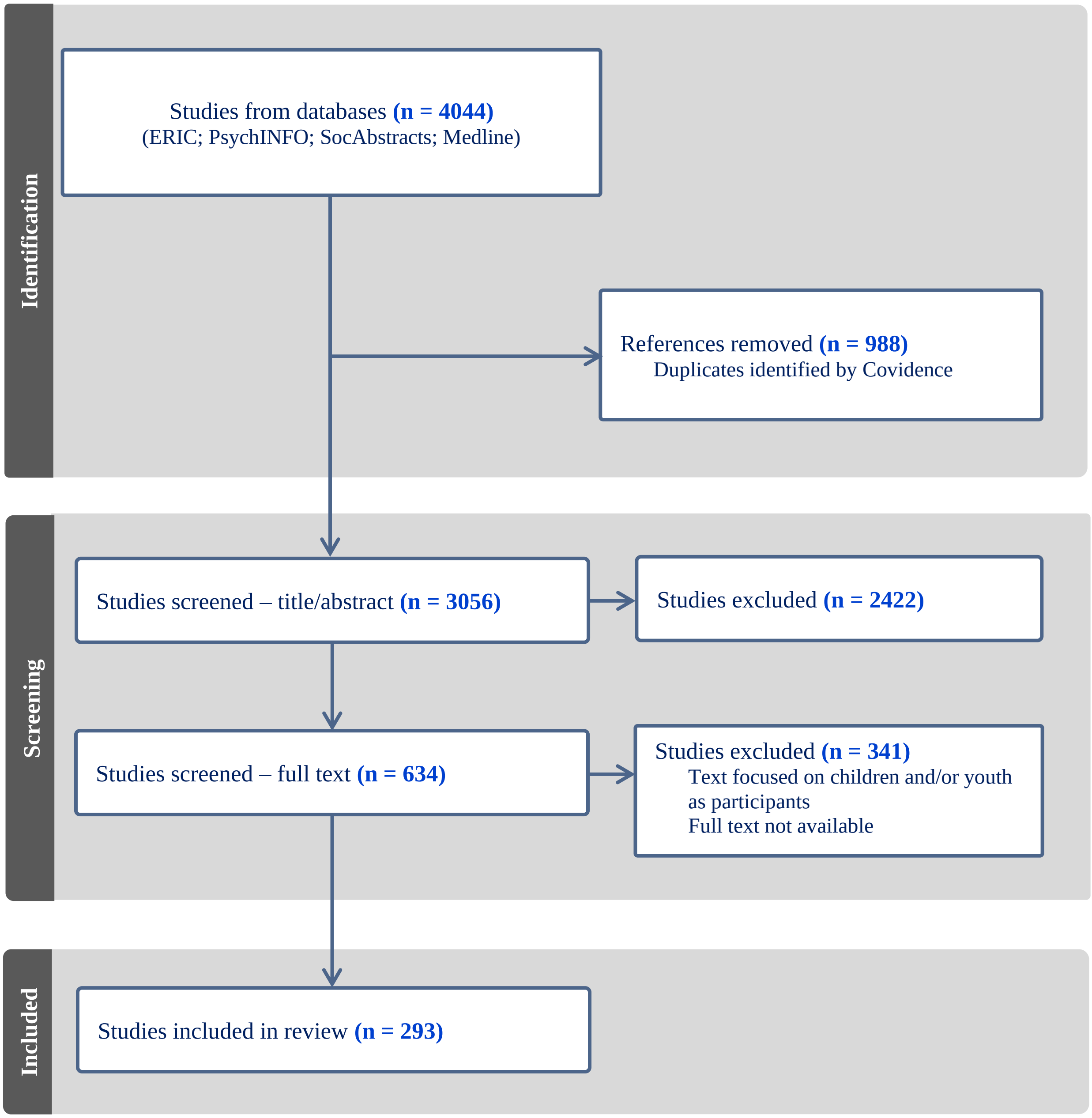
