## Appendix C for "More Than Words: An Integrative Review of Innovative Elicitation Techniques for Qualitative Interviews"

### Appendix C: Elicitation Techniques - Categorized and Sorted by Author

| Authors | Title | Published Year | Journal | Volume | Issue | Pages | DOI | Technique(s) |
|---|---|---|---|---|---|---|---|---|
| Affleck, William; Glass, K. C; MacDonald, Mary Ellen | The limitations of language: Male participants, stoicism, and the qualitative research interview. | 2013 | American Journal of Men's Health | 7 | 2 | 155-162 | http://dx.doi.org/10.1177/1557988312464038 | photo-elicitation |
| Aldridge, Jo | Working with vulnerable groups in social research: dilemmas by default and design | 2014 | Qualitative Research | 14 | 1 | 112-130 | http://dx.doi.org/10.1177/1468794112455041 | photo-elicitation |
| Allen, Quaylan | Photographs and stories: ethics, benefits and dilemmas of using participant photography with Black middle-class male youth | 2012 | Qualitative Research | 12 | 4 | 443-458 | http://dx.doi.org/10.1177/1468794111433088 | photo-elicitation |
| Anderson, Terilynn | Photovoice as a catalyst for conversation: Children as co-researchers in an elementary school in the USA. | 2015 | Photography in educational research: Critical reflections from diverse contexts. | | | 36-49 | | photo-elicitation |
| Antelius, Eleonor; Kiwi, Mahin; Strandroos, Lisa | Ethnographic methods for understanding practices around dementia among culturally and linguistically diverse people. | 2018 | Social research methods in dementia studies: Inclusion and innovation. | | | 121-139 | | reflection |
| Ashleigh, Melanie J; Meyer, Edgar | Deepening the understanding of trust: Combining repertory grid and narrative to explore the uniqueness of trust. | 2012 | Handbook of research methods on trust. | | | 138-148 | | repertory grid |
| Ayala, Ricardo A; Koch, Tomas F | The Image of Ethnographyâ€"Making Sense of the Social Through Images: A Structured Method | 2019 | International Journal of Qualitative Methods | 18 | | | http://dx.doi.org/10.1177/1609406919843014 | drawing |
| Babapour, Maral; Rehammar, Bjorn; Rahe, Ulrike | A Comparison of Diary Method Variations for Enlightening Form Generation in the Design Process | 2012 | Design and Technology Education | 17 | 3 | 49-60 | | diary |
| Baker, Amanda A; Lee, Joseph J | Mind the Gap: Unexpected Pitfalls in Doing Classroom Research | 2011 | The Qualitative Report | 16 | 5 | 1435-1447 | | stimulated recall |
| Balbale, Salva Najib; Schwingel, Andiara; Chodzko-Zajko, Wojtek; Huhman, Marian | Visual and participatory research methods for the development of health messages for underserved populations. | 2014 | Health Communication | 29 | 7 | 728-740 | http://dx.doi.org/10.1080/10410236.2013.800442 | photo-elicitation |
| Balmer, Claire; Griffiths, Frances; Dunn, Janet | A 'new normal': Exploring the disruption of a poor prognostic cancer diagnosis using interviews and participant-produced photographs | 2015 | Health | 19 | 5 | 451-472 | http://dx.doi.org/10.1177/1363459314554319 | photo-elicitation |
| Balmer, Claire; Griffiths, Frances; Dunn, Janet | A review of the issues and challenges involved in using participant-produced photographs in nursing research. | 2015 | Journal of Advanced Nursing | 71 | 7 | 1726-1737 | http://dx.doi.org/10.1111/jan.12627 | photo-elicitation |
| Balomenou, Nika; Garrod, Brian | A Review of Participant-Generated Image Methods in the Social Sciences | 2016 | Journal of Mixed Methods Research | 10 | 4 | 335-351 | http://dx.doi.org/10.1177/1558689815581561 | photo-elicitation |
| Barbeiro, Ana; Spini, Dario | Calendar interviewing: A mixed methods device for a biographical approach to migration. | 2017 | Qualitative Research in Psychology | 14 | 1 | 81-107 | http://dx.doi.org/10.1080/14780887.2016.1249581 | life grid |
| Barrantes-Elizondo, Lena | Creating Space for Visual Ethnography in Educational Research | 2019 | Online Submission | 23 | 2 | | | photo-elicitation; drawing |
| Bartlett, Ruth | Modifying the diary interview method to research the lives of people with dementia | 2012 | Qualitative Health Research | 22 | 12 | 1717-1726 | http://dx.doi.org/10.1177/1049732312462240 | diary |
| Barton, Keith C. | Elicitation Techniques: Getting People to Talk about Ideas They Don't Usually Talk About | 2015 | Theory and Research in Social Education | 43 | 2 | 179-205 | http://dx.doi.org/10.1080/00933104.2015.1034392 | Review across categories |
| Bates, Elizabeth A; McCann, Joseph J; Kaye, Linda K; Taylor, Julie C | "Beyond words": A researcher's guide to using photo elicitation in psychology. | 2017 | Qualitative Research in Psychology | 14 | 4 | 459-481 | http://dx.doi.org/10.1080/14780887.2017.1359352 | photo-elicitation |
| Beaudin, Bennett; Maar, Marion; Manitowabi Darrel; Moeke-Pickering Taima; Trudeau-Peltier Doreen; Trudeau, Sheila | The Gaataaâ€™aabing Visual Research Method: A Culturally Safe Anishinaabek Transformation of Photovoice | 2019 | International Journal of Qualitative Methods | 18 | | | http://dx.doi.org/10.1177/1609406919851635 | photo-elicitation |
| Becker, Larissa | Methodological proposals for the study of consumer experience | 2018 | Qualitative Market Research: An International Journal | 21 | 4 | 465-490 | http://dx.doi.org/10.1108/QMR-01-2017-0036 | diary |
| Bendell, Katherine; Sylvestre, John | How different approaches to taking pictures influences participation in a photovoice project | 2017 | Action Research | 15 | 4 | 357-372 | http://dx.doi.org/10.1177/1476750316653812 | photo-elicitation |
| Bendixen, Lisa D.; Klimow, Nicole | Participatory Concept Mapping as an Integration Tool in Mixed Methods Research: Exploring Preservice Teachers' Epistemic Cognition and Teaching Orientation | 2019 | International Journal of Educational Methodology | 5 | 2 | 247-264 | | concept mapping |
| Benedetti, Allison; Jackson, John; Luo, Lili | Vignettes: Implications for LIS Research | 2018 | College & Research Libraries | 79 | 2 | 222-236 | | vignette |
| Berends, Lynda | Embracing the Visual: Using Timelines with In-Depth Interviews on Substance Use and Treatment | 2011 | Qualitative Report | 16 | 1 | | | timeline |
| Berg, Tessa; Bowen, Tracey; Smith, Colin; Smith, Sally | Visualising the Future: Surfacing Student Perspectives on Post-Graduation Prospects Using Rich Pictures | 2017 | Higher Education Research and Development | 36 | 7 | 1339-1354 | http://dx.doi.org/10.1080/07294360.2017.1325855 | rich picture |

| | | | | | | | | |
|---|---|---|---|---|---|---|---|---|
| Bessette, Harriet J.; Paris, Nita A. | Using Visual and Textual Metaphors to Explore Teachers' Professional Roles and Identities | 2020 | International Journal of Research & Method in Education | 43 | 2 | 173-188 | https://doi.org/10.1080/1743727X.2019.1611759 | drawing |
| Birkeland, Asta | Research Dilemmas Associated with Photo Elicitation in Comparative Early Childhood Education Research | 2013 | Research in Comparative and International Education | 8 | 4 | 455-467 | http://dx.doi.org/10.2304/rcie.2013.8.4.455 | photo-elicitation |
| Blikstad-Balas, Marte | Key Challenges of Using Video When Investigating Social Practices in Education: Contextualization, Magnification, and Representation | 2017 | International Journal of Research & Method in Education | 40 | 5 | 511-523 | https://doi.org/10.1080/1743727X.2016.1181162 | video-elicitation |
| Bloch, Stefano | Place-based elicitation: Interviewing graffiti writers at the scene of the crime. | 2018 | Journal of Contemporary Ethnography | 47 | 2 | 171-198 | http://dx.doi.org/10.1177/0891241616639640 | place-elicitation |
| Boden, Zoe; Larkin, Michael; Iyer, Malvika | Picturing ourselves in the world: Drawings, interpretative phenomenological analysis and the relational mapping interview. | 2019 | Qualitative Research in Psychology | 16 | 2 | 218-236 | http://dx.doi.org/10.1080/14780887.2018.1540679 | mapping |
| Bonnycastle, Marleny M.; Bonnycastle, Colin R. | Photographs Generate Knowledge: Reflections on Experiential Learning In/Outside the Social Work Classroom | 2015 | Journal of Teaching in Social Work | 35 | 3 | 233-250 | http://dx.doi.org/10.1080/08841233.2015.1027031 | photo-elicitation |
| Bowes-Catton, Helen; Barker, Meg; Richards, Christina | I didn't know that I could feel this relaxed in my body': Using visual methods to research bisexual people's embodied experiences of identity and space. | 2011 | Visual methods in psychology: Using and interpreting images in qualitative research. | | | 255-270 | | photo-elicitation; diary |
| Bowleg, Lisa | Towards a Critical Health Equity Research Stance: Why Epistemology and Methodology Matter More than Qualitative Methods | 2017 | Health Education & Behavior | 44 | 5 | 677-684 | http://dx.doi.org/10.1177/1090198117728760 | photo-elicitation |
| Boxall, Kathy; Ralph, Sue | Research Ethics Committees and the Benefits of Involving People with Profound and Multiple Learning Disabilities in Research | 2011 | British Journal of Learning Disabilities | 39 | 3 | 173-180 | http://dx.doi.org/10.1111/j.1468-3156.2010.00645.x | photo-elicitation |
| Bradbury-Jones, C; Taylor, J; Herber, OR | Vignette development and administration: a framework for protecting research participants | 2014 | International Journal of Social Research Methodology | 17 | 4 | 427-440 | http://dx.doi.org/10.1080/13645579.2012.750833 | vignette |
| Branch, Nicole A. | Illuminating Social Justice in the Framework: Transformative Methodology, Concept Mapping and Learning Outcomes Development for Critical Information Literacy | 2019 | Communications in Information Literacy | 13 | 1 | | | concept mapping |
| Bravington Alison; King, Nigel | Putting graphic elicitation into practice: tools and typologies for the use of participant-led diagrams in qualitative research interviews | 2019 | Qualitative Research | 19 | 5 | 506-523 | http://dx.doi.org/10.1177/1468794118781718 | diagram |
| Breen, Andrea V; Scott, Christine; McLean, Kate C | The "stuff" of narrative identity: Touring big and small stories in emerging adults' dorm rooms. | 2019 | Qualitative Psychology | | | | http://dx.doi.org/10.1037/qup0000158 | life grid |
| Brooks, Rachel; Lainio, Anu; Lažetić, Predrag | Using creative methods to research across difference. An introduction to the special issue, International Journal of Social Research Methodology, 23:1, 1-6, DOI: 10.1080/13645579.2019.1672281 | 2020 | International journal of social research methodology | 23 | 1 | 1-6 | https://doi.org/10.1080/13645579.2019.1672281 | Review across categories |
| Brown, Kyla; Worrall, Linda; Davidson, Bronwyn; Howe, Tami | Reflection on the benefits and limitations of participant-generated photography as an adjunct to qualitative interviews with participants with aphasia. | 2013 | Aphasiology | 27 | 10 | 1214-1231 | http://dx.doi.org/10.1080/02687038.2013.808736 | photo-elicitation |
| Brown, Nicole | Identity boxes: using materials and metaphors to elicit experiences | 2019 | International Journal of Social Research Methodology | 22 | 5 | 487-501 | http://dx.doi.org/10.1080/13645579.2019.1590894 | object-elicitation |
| Brushwood Rose, Chloe; Granger, Colette A. | Unexpected Self-Expression and the Limits of Narrative Inquiry: Exploring Unconscious Dynamics in a Community-Based Digital Storytelling Workshop | 2013 | International Journal of Qualitative Studies in Education (QSE) | 26 | 2 | 216-237 | https://doi.org/10.1080/09518398.2012.666286 | story-telling |
| Bryanton Olive; Weeks, Lori; Townsend, Elizabeth; Montelpare, William; Lees, Jessie; Moffatt Lyndsay | The Utilization and Adaption of Photovoice With Rural Women Aged 85 and Older | 2019 | International Journal of Qualitative Methods | 18 | | | http://dx.doi.org/10.1177/1609406919883450 | photo-elicitation |
| Bugos, Eva; Frasso, Rosemary; FitzGerald, Elizabeth; True, Gala; Adachi-Mejia, Anna M; Cannuscio, Carolyn | Practical guidance and ethical considerations for studies using photo-elicitation interviews. | 2014 | Preventing Chronic Disease: Public Health Research, Practice, and Policy | 11 | | | | photo-elicitation |
| Burgess-Allen, Jilla; Owen-Smith, Vicci | Using mind mapping techniques for rapid qualitative data analysis in public participation processes. | 2010 | Health Expectations: An International Journal of Public Participation in Health Care & Health Policy | 13 | 4 | 406-415 | http://dx.doi.org/10.1111/j.1369-7625.2010.00594.x | concept mapping; mind mapping |
| Burkart, Thomas; Weggen, Jenny | Dialogic introspection: A method for exploring emotions in everyday life and experimental contexts. | 2015 | Methods of exploring emotions. | | | 101-111 | | reflection, dialogic introspection |
| Burke, Dawn; Evans, Joan | Embracing the Creative: The Role of Photo Novella in Qualitative Nursing Research | 2011 | International Journal of Qualitative Methods | 10 | 2 | 164-177 | http://dx.doi.org/10.1177/160940691101000205 | photo-elicitation |
| Burles Meridith; Thomas, Roanne | â€œI Just Don't Think There's any other Image that Tells the Story like [This] Picture Doesâ€: Researcher and Participant Reflections on the Use of Participant-Employed Photography in Social Research | 2014 | International Journal of Qualitative Methods | 13 | 1 | 185-205 | http://dx.doi.org/10.1177/160940691401300107 | photo-elicitation |

| | | | | | | | | |
|---|---|---|---|---|---|---|---|---|
| Burton, Amy; Hughes, Melanie; Dempsey, Robert C | Quality of life research: A case for combining photo-elicitation with interpretative phenomenological analysis. | 2017 | Qualitative Research in Psychology | 14 | 4 | 375-393 | http://dx.doi.org/10.1080/14780887.2017.1322650 | photo-elicitation |
| Campbell, Sarah; Ward, Richard | Video and observation data as a method to document practice and performances of gender in the dementia care-based hair salon: Practices and processes. | 2018 | Social research methods in dementia studies: Inclusion and innovation. | | | 96-117 | | video-elicitation |
| Capous-Desyllas Moshoula; Bromfield, Nicole F | Using an Arts-Informed Eclectic Approach to Photovoice Data Analysis | 2018 | International Journal of Qualitative Methods | 17 | 1 | | http://dx.doi.org/10.1177/1609406917752189 | photo-elicitation |
| Carroll, Katherine; Mesman, Jessica | Multiple researcher roles in video-reflexive ethnography. | 2018 | Qualitative Health Research | 28 | 7 | 1145-1156 | http://dx.doi.org/10.1177/1049732318759490 | video-elicitation |
| Cassidy, Angela; Maule, John | Risk communication and participatory research: 'Fuzzy felt', visual games and group discussion of complex issues. | 2011 | Visual methods in psychology: Using and interpreting images in qualitative research. | | | 205-222 | | fuzzy felt |
| Catalani, Caricia; Minkler, Meredith | Photovoice: A Review of the Literature in Health and Public Health | 2010 | Health Education & Behavior | 37 | 3 | 424-451 | https://doi.org/10.1177/1090198109342084 | photo-elicitation; photovoice |
| Catney, Gemma; Frost, Diane; Vaughn, Leona | Residents' perspectives on defining neighbourhood: Mental mapping as a tool for participatory neighbourhood research. | 2019 | Qualitative Research | 19 | 6 | 735-752 | http://dx.doi.org/10.1177/1468794118803841 | mapping |
| Chang, Jamie Suki | The Docent Method: A Grounded Theory Approach for Researching Place and Health. | 2017 | Qualitative health research | 27 | 4 | 609-619 | https://dx.doi.org/10.1177/1049732316667055 | walking |
| Chawla-Duggan, Rita; Konantambigi, Rajani; Lam, Michelle Mei Seung; Sollied, Sissel | A visual methods approach for researching children's perspectives: capturing the dialectic and visual reflexivity in a cross-national study of father-child interactions | 2020 | International Journal of Social Research Methodology | 23 | 1 | 37-54 | http://dx.doi.org/10.1080/13645579.2019.1672283 | video-elicitation |
| Chawla-Duggan, Rita; Milner, Susan; Porter, Jill | Reflexivity and visual technology in research: young childrenâ€™s perspectives of paternal engagement in the home environment | 2018 | Qualitative Research | 18 | 4 | 471-491 | http://dx.doi.org/10.1177/1468794117728412 | video-elicitation |
| Chen, Annie T | Timeline Drawing and the Online Scrapbook | 2018 | International Journal of Qualitative Methods | 17 | 1 | | http://dx.doi.org/10.1177/1609406917753207 | timeline; collage |
| Chen, Julienne; Neo Pearlyn | Texting the waters: An assessment of focus groups conducted via the WhatsApp smartphone messaging application | 2019 | Methodological Innovations | 12 | 3 | | http://dx.doi.org/10.1177/2059799119884276 | card task |
| Ciolan Lucian; Manasia Loredana | Reframing Photovoice to Boost Its Potential for Learning Research | 2017 | International Journal of Qualitative Methods | 16 | 1 | | http://dx.doi.org/10.1177/1609406917702909 | photo-elicitation |
| Clark, Alison | Breaking Methodological Boundaries? Exploring Visual, Participatory Methods with Adults and Young Children | 2011 | European Early Childhood Education Research Journal | 19 | 3 | 321-330 | http://dx.doi.org/10.1080/1350293X.2011.597964 | photo-elicitation; mapping |
| Clark, Andrew; Morriss, Lisa | The use of visual methodologies in social work research over the last decade: A narrative review and some questions for the future | 2017 | Qualitative Social Work | 16 | 1 | 29-43 | http://dx.doi.org/10.1177/1473325015601205 | mapping; photo-elicitation |
| Cluley, Victoria | Using Photovoice to Include People with Profound and Multiple Learning Disabilities in Inclusive Research | 2017 | British Journal of Learning Disabilities | 45 | 1 | 39-46 | http://dx.doi.org/10.1111/bld.12174 | photo-elicitation |
| Coemans Sara; Anne-Leen, Raymakers; Vandenabeele Joke; Hannes, Karin | Evaluating the extent to which social researchers apply feminist and empowerment frameworks in photovoice studies with female participants: A literature review | 2019 | Qualitative Social Work | 18 | 1 | 37-59 | http://dx.doi.org/10.1177/1473325017699263 | photo-elicitation |
| CohenMiller, Anna S | Visual Arts as a Tool for Phenomenology | 2018 | Forum : Qualitative Social Research | 19 | 1 | | | drawing |
| Collier, Aileen; Wyer, Mary | Researching reflexively with patients and families: Two studies using video-reflexive ethnography to collaborate with patients and families in patient safety research. | 2016 | Qualitative Health Research | 26 | 7 | 979-993 | http://dx.doi.org/10.1177/1049732315618937 | video-elicitation |
| Consuegra, Els; Engels, Nadine; Willegems, Vicky | Using video-stimulated recall to investigate teacher awareness of explicit and implicit gendered thoughts on classroom interactions | 2016 | Teachers and Teaching: Theory and Practice | 22 | 6 | 683-699 | http://dx.doi.org/10.1080/13540602.2016.1158958 | stimulated recall |
| Cooper, Cheryl M; Yarbrough, Susan P | Tell me-Show me: Using combined focus group and photovoice methods to gain understanding of health issues in rural Guatemala. | 2010 | Qualitative Health Research | 20 | 5 | 644-653 | http://dx.doi.org/10.1177/1049732310361894 | photo-elicitation |
| Cooper, Cheryl; Sorensen, William; Yarbrough, Susan | Visualising the Health of Communities: Using Photovoice as a Pedagogical Tool in the College Classroom | 2017 | Health Education Journal | 76 | 4 | 454-466 | http://dx.doi.org/10.1177/0017896917691790 | photo-elicitation |
| Copeland, Andrea J; Agosto, Denise E | Diagrams and Relational Maps: The Use of Graphic Elicitation Techniques with Interviewing for Data Collection, Analysis, and Display | 2012 | International Journal of Qualitative Methods | 11 | 5 | 513-533 | http://dx.doi.org/10.1177/160940691201100501 | mapping |
| Copes, Heith; Tchoula, Whitney; Brookman, Fiona; Ragland, Jared | Photo-Elicitation Interviews with Vulnerable Populations: Practical and Ethical Considerations | 2018 | Deviant Behavior | 39 | 4 | 475-494 | http://dx.doi.org/10.1080/01639625.2017.1407109 | photo-elicitation |
| Cox, Andrew; Benson, Melanie | Visual Methods and Quality in Information Behaviour Research: The Cases of Photovoice and Mental Mapping | 2017 | Information Research: An International Electronic Journal | 22 | 2 | | | photo-elicitation |
| Cudworth Erika | Now, where were we?: The highs and lows of hunting data with a research pack | 2018 | Journal of Sociology | 54 | 4 | 488-503 | http://dx.doi.org/10.1177/1440783318816761 | walking |
| D'angelo Alessio; Ryan, Louise; Tubaro Paola | Visualization in Mixed-Methods Research on Social Networks | 2016 | Sociological Research Online | 21 | 2 | 1-4 | http://dx.doi.org/10.5153/sro.3996 | photo-elicitation |
| Daniel, Leah | A methodology to investigate pre-service teachersâ€™ content-related instructional decisions and teaching actions | 2018 | Asia-Pacific Journal of Teacher Education | 46 | 5 | 478-494 | http://dx.doi.org/10.1080/1359866X.2018.1444139 | stimulated recall |

| | | | | | | | | |
|---|---|---|---|---|---|---|---|---|
| Darnhofer Ika | Using Comic-Style Posters for Engaging Participants and for Promoting Researcher Reflexivity | 2018 | International Journal of Qualitative Methods | 17 | 1 | | http://dx.doi.org/10.1177/1609406918804716 | comic |
| Davies, Simon J; Bourke, Lorna J | Crossing over cultures: Using visual methods in a cross-cultural context for teaching and research. | 2017 | Teaching visual methods in the social sciences. | | | 162-181 | | photo-elicitation; drawing |
| Day, Melissa; Thatcher, Joanne | "I'm really embarrassed that you're going to read this ...": Reflections on using diaries in qualitative research. | 2009 | Qualitative Research in Psychology | 6 | 4 | 249-259 | http://dx.doi.org/10.1080/14780880802070583 | diary |
| de Groot, Tjitske; Jacquet, Wolfgang; Free De Backer; Peters, Ruth; Meurs, Pieter | Using visual vignettes to explore sensitive topics: a research note on exploring attitudes towards people with albinism in Tanzania | 2020 | International Journal of Social Research Methodology | 23 | 6 | 749-755 | http://dx.doi.org/10.1080/13645579.2020.1757250 | vignette; photo-elicitation |
| de Jager, AdÃ¨le; Fogarty, Andrea; Tewson, Anna; Lenette, Caroline; Boydell, Katherine M | Digital Storytelling in Research: A Systematic Review | 2017 | The Qualitative Report | 22 | 10 | 2548-2582 | | story-telling |
| de Souza Daniela Maysa; Backes VÃ¢nia Marli Schubert; do Prado Marta Lenise; Martini Jussara Gue; Medina Moya JosÃ© Luis | The Use of Autoscopy From the Epistemological Perspective of Action Research for Self-Analysis and Reflection of Teacher Practice | 2019 | International Journal of Qualitative Methods | 18 | | | http://dx.doi.org/10.1177/1609406919873247 | video-elicitation |
| Del Busso, Lilliana | Using photographs to explore the embodiment of pleasure in everyday life. | 2011 | Visual methods in psychology: Using and interpreting images in qualitative research. | | | 43-54 | | vignette |
| Dew, Angela; Smith, Louisa; Collings, Susan; Isabella Dillon Savage | Complexity Embodied: Using Body Mapping to Understand Complex Support Needs | 2018 | Forum : Qualitative Social Research | 19 | 2 | | | body mapping |
| Durso, Francis T; Kazi, Sadaf; Ferguson, Ashley N | The Threat-Strategy Interview. | 2015 | Applied Ergonomics | 47 | | 336-344 | http://dx.doi.org/10.1016/j.apergo.2014.08.001 | situation-elicitation |
| Dutta Uttaran | Conducting Ethnographic Research in Low Literate, Economically Weak Underserved Spaces: An Introduction to Iconic Legisigns-Guided Interviewing (ILGI) | 2019 | International Journal of Qualitative Methods | 18 | | | http://dx.doi.org/10.1177/1609406919855279 | object-elicitation |
| Dutton, S; Davison, C M; Malla, M; Bartels, S; Collier, K; Plamondon, K; Purkey, E | Biographical Collage as a Tool in Inuit Community-Based Participatory Research and Capacity Development | 2019 | International Journal of Qualitative Methods | 18 | | | http://dx.doi.org/10.1177/1609406919877307 | collage |
| Elliot, Dely Lazarte; Reid, Kate; Baumfield, Vivienne | Capturing Visual Metaphors and Tales: Innovative or Elusive? | 2017 | International Journal of Research & Method in Education | 40 | 5 | 480-496 | http://dx.doi.org/10.1080/1743727X.2016.1181164 | photo-elicitation |
| Erdner, Anette; Magnusson, Annabella | Photography as a method of data collection: Helping people with long-term mental illness to convey their life world. | 2011 | Perspectives in Psychiatric Care | 47 | 3 | 145-150 | http://dx.doi.org/10.1111/j.1744-6163.2010.00283.x | photo-elicitation |
| Esin, Cigdem; Squire, Corinne | Visual Autobiographies in East London: Narratives of Still Images, Interpersonal Exchanges, and Intrapersonal Dialogues | 2013 | Forum Qualitative Sozialforschung/Forum: Qualitative Social Research | 14 | 2 | | | drawing; collage |
| Evans-Agnew, Robin A; Rosemberg, Marie-Anne S | Questioning photovoice research: Whose voice? | 2016 | Qualitative Health Research | 26 | 8 | 1019-1030 | http://dx.doi.org/10.1177/1049732315624223 | photo-elicitation |
| Fang, Mei Lan; Sixsmith, Judith; Woolrych, Ryan; Canham, Sarah L; Battersby, Lupin; Ren, Tori Hui; Sixsmith, Andrew | Integrating sense of place within new housing developments: A community-based participatory research approach. | 2018 | Resilience and ageing: Creativity, culture and community. | | | 129-156 | http://dx.doi.org/10.1332/policypress/9781447340911.003.0007 | photo-elicitation |
| Fayette, Rainart; Bond, Caroline | A Systematic Literature Review of Qualitative Research Methods for Eliciting the Views of Young People with ASD about Their Educational Experiences | 2018 | European Journal of Special Needs Education | 33 | 3 | 349-365 | http://dx.doi.org/10.1080/08856257.2017.1314111 | Review across categories |
| Fearon Kriss | â€˜Have you ever talked to any women with Turner syndrome?â€™ Using universal design and photo elicitation interviews in research with women with mild cognitive impairment | 2019 | Methodological Innovations | 12 | 2 | | http://dx.doi.org/10.1177/2059799119841933 | photo-elicitation |
| Felsher, Marisa; Wiehe, Sarah E.; Gunn, Jayleen K. L.; Roth, Alexis M. | â€˜I got it off my chestâ€™: An examination of how research participation improved the mental health of women engaging in transactional sex | 2018 | Community Mental Health Journal | 54 | 2 | 171-179 | http://dx.doi.org/10.1007/s10597-017-0094-y | diary |
| Filep, Crystal Victoria; Turner, Sarah; Eidse, Noelani; Thompson-Fawcett, Michelle; Fitzsimons, Sean | Advancing rigour in solicited diary research | 2018 | Qualitative Research | 18 | 4 | 451-470 | http://dx.doi.org/10.1177/1468794117728411 | diary |
| Fitt, Helen | Researching mobile practices: Participant reflection and audio-recording in Repeat Question Diaries | 2018 | Qualitative Research | 18 | 6 | 654-670 | http://dx.doi.org/10.1177/1468794117743462 | diary |
| Frith, Hannah | Narrating biographical disruption and repair: Exploring the place of absent images in women's experiences of cancer and chemotherapy. | 2011 | Visual methods in psychology: Using and interpreting images in qualitative research. | | | 55-68 | | photo-elicitation |
| Fritz, Heather A. | Learning to do better: The transactional model of diabetes self-management integration | 2015 | Qualitative Health Research | 25 | 7 | 875-886 | http://dx.doi.org/10.1177/1049732314552453 | photo-elicitation; diary |

| | | | | | | | | |
|---|---|---|---|---|---|---|---|---|
| Fritz, Roschelle L; Vandermause, Roxanne | Data collection via in-depth email interviewing: Lessons from the field. | 2018 | Qualitative Health Research | 28 | 10 | 1640-1649 | http://dx.doi.org/10.1177/1049732316689067 | writing (email) |
| Fullana, Judit; Pallisera, Maria; Vila, Montserrat | Advancing towards inclusive social research: visual methods as opportunities for people with severe mental illness to participate in research | 2014 | International Journal of Social Research Methodology | 17 | 6 | 723-738 | http://dx.doi.org/10.1080/13645579.2013.832049 | photo-elicitation; drawing |
| Futch, Valerie A; Fine, Michelle | Mapping as a method: History and theoretical commitments. | 2014 | Qualitative Research in Psychology | 11 | 1 | 42-59 | http://dx.doi.org/10.1080/14780887.2012.719070 | mapping |
| Galman, Sally A. C | The truthful messenger: Visual methods and representation in qualitative research in education. | 2009 | Qualitative Research | 9 | 2 | 197-217 | http://dx.doi.org/10.1177/1468794108099321 | drawing |
| GarcÃ-a, Borja; Welford, Jo; Smith, Brett | Using a smartphone app in qualitative research: the good, the bad and the ugly | 2016 | Qualitative Research | 16 | 5 | 508-525 | http://dx.doi.org/10.1177/1468794115593335 | photo-elicitation; diary |
| Gastaldo, Denise; Rivas-Quarneti, Natalia; Magalhaes, Lilian | Body-Map Storytelling as a Health Research Methodology: Blurred Lines Creating Clear Pictures | 2018 | Forum : Qualitative Social Research | 19 | 2 | | | body mapping |
| Genoe, M. Rebecca; Dupuis, Sherry L. | Picturing Leisure: Using Photovoice to Understand the Experience of Leisure and Dementia | 2013 | Qualitative Report | 18 | | | | photo-elicitation |
| Glass, Christine | "There's Not Much Room for Anything to Go Amiss": Narrative and Arts-Based Inquiry in Teacher Education | 2011 | Issues in Educational Research | 21 | 2 | 130-144 | | drawing |
| Glaw Xanthe; Inder, Kerry; Kable Ashley; Hazelton, Michael | Visual Methodologies in Qualitative Research | 2017 | International Journal of Qualitative Methods | 16 | 1 | | http://dx.doi.org/10.1177/1609406917748215 | photo-elicitation |
| Glenis, Mark; Boulton, Amohia | Indigenising Photovoice: Putting MÄori Cultural Values Into a Research Method | 2017 | Forum : Qualitative Social Research | 18 | 3 | | | photo-elicitation |
| Goldman, Ellen F; Swayze, Susan | In-Depth Interviewing with Healthcare Corporate Elites: Strategies for Entry and Engagement | 2012 | International Journal of Qualitative Methods | 11 | 3 | 230-243 | http://dx.doi.org/10.1177/160940691201100304 | concept mapping |
| Gonzales, Leslie D.; Rincones, Rodolfo | Using Participatory Action Research and Photo Methods to Explore Higher Education Administration as an Emotional Endeavor | 2013 | Qualitative Report | 18 | | | | photo-elicitation |
| Goopy Suzanne; Kassan Anusha | Arts-Based Engagement Ethnography: An Approach for Making Research Engaging and Knowledge Transferable When Working With Harder-to-Reach Communities | 2019 | International Journal of Qualitative Methods | 18 | | | http://dx.doi.org/10.1177/1609406918820424 | object-elicitation |
| Grellier, Jane | Rhizomatic mapping: Spaces for learning in higher education. | 2013 | Higher Education Research & Development | 32 | 1 | 83-95 | http://dx.doi.org/10.1080/07294360.2012.750280 | mapping |
| Gubrium, Aline; Harper, Krista | Participatory visual and digital methods. | 2013 | Routledge | | | | | photo-elicitation; video-elicitation; story-telling; mapping |
| Guell, Cornelia; Ogilvie, David | Picturing commuting: Photovoice and seeking well-being in everyday travel. | 2015 | Qualitative Research | 15 | 2 | 201-218 | http://dx.doi.org/10.1177/1468794112468472 | photo-elicitation |
| Hadfield, Mark; Haw, Kaye | Video: Modalities and Methodologies | 2012 | International Journal of Research & Method in Education | 35 | 3 | 311-324 | https://doi.org/10.1080/1743727X.2012.717434 | video-elicitation |
| Hall, Pamela D.; Bowen, Glenn A. | The Use of Photovoice for Exploring Students' Perspectives on Themselves and Others | 2015 | Journal of Ethnographic & Qualitative Research | 9 | 3 | 196-208 | | photo-elicitation; photovoice |
| Han, Christina S; Oliffe, John L | Photovoice in mental illness research: A review and recommendations. | 2016 | Health: An Interdisciplinary Journal for the Social Study of Health, Illness and Medicine | 20 | 2 | 110-126 | http://dx.doi.org/10.1177/1363459314567790 | photo-elicitation |
| Harley, Anne | Picturing Reality: Power, Ethics, and Politics in Using Photovoice | 2012 | International Journal of Qualitative Methods | 11 | 4 | 320-339 | http://dx.doi.org/10.1177/160940691201100402 | photo-elicitation |
| Harris, Magdalena; Rhodes, Tim | "It's not much of a life": The benefits and ethics of using life history methods with people who inject drugs in qualitative harm reduction research. | 2018 | Qualitative Health Research | 28 | 7 | 1123-1134 | http://dx.doi.org/10.1177/1049732318764393 | timeline; life grid |
| Hartel Jenna; Noone, Rebecca; Oh, Christie; Power, Stephanie; Danzanov Pavel; Kelly, Bridgette | The iSquare protocol: combining research, art, and pedagogy through the draw-and-write technique | 2018 | Qualitative Research | 18 | 4 | 433-450 | http://dx.doi.org/10.1177/1468794117722193 | drawing |
| Harvey, Laura | Intimate reflections: Private diaries in qualitative research | 2011 | Qualitative Research | 11 | 6 | 664-682 | http://dx.doi.org/10.1177/1468794111415959 | diary |
| Hegarty, Ann | Photovoice: Facilitating Fathers' Narratives of Care | 2016 | Adult Learner: The Irish Journal of Adult and Community Education | | | | | photo-elicitation |
| Hellzen, Ove; Haugenes, Marit; Ã˜stby, May | â€˜Itâ€™s my home and your workâ€™: The views of a filmed vignette describing a challenging everyday situation from the perspective of people with intellectual disabilities | 2018 | International Journal of Qualitative Studies on Health and Well-being | 13 | 1 | 9 | http://dx.doi.org/10.1080/17482631.2018.1468198 | vignette |
| Henry, Stephen G.; Fetters, Michael D. | Video elicitation interviews: A qualitative research method for investigating physician-patient interactions | 2012 | Annals of Family Medicine | 10 | 2 | 118-125 | http://dx.doi.org/10.1370/afm.1339 | video-elicitation |
| Herbst, Patricio; Chazan, Daniel | Studying Professional Knowledge Use in Practice Using Multimedia Scenarios Delivered Online | 2015 | International Journal of Research & Method in Education | 38 | 3 | 272-287 | http://dx.doi.org/10.1080/1743727X.2015.1025742 | storyboard; cartoons |

| | | | | | | | | |
|---|---|---|---|---|---|---|---|---|
| Hergenrather, Kenneth C; Rhodes, Scott D; Cowan, Chris A; Bardhoshi, Gerta; Pula, Sara | Photovoice as community-based participatory research: A qualitative review. | 2009 | American Journal of Health Behavior | 33 | 6 | 686-698 | http://dx.doi.org/10.5993/AJHB.33.6.6 | photo-elicitation |
| Hermanns, Melinda; Greer, Danice B; Cooper, Cheryl | Visions of Living with Parkinson's Disease: A Photovoice Study | 2015 | The Qualitative Report | 20 | 3 | 336-355 | | photo-elicitation |
| Herron, Rachel; Dansereau, Lisette; Wrathall, Meghan; Funk, Laura; Spencer, Dale | Using a flexible diary method rigorously and sensitively with family carers | 2019 | Qualitative Health Research | 29 | 7 | 1004-1015 | http://dx.doi.org/10.1177/1049732318816081 | diary |
| Higgins, Marc | Placing Photovoice under Erasure: A Critical and Complicit Engagement with What It Theoretically Is (Not) | 2016 | International Journal of Qualitative Studies in Education (QSE) | 29 | 5 | 670-685 | http://dx.doi.org/10.1080/09518398.2016.1145276 | photo-elicitation |
| Higgins, Marc | Rebraiding Photovoice: Methodological MÃ©tissage at the Cultural Interface | 2014 | Australian Journal of Indigenous Education | 43 | 2 | 208-217 | http://dx.doi.org/10.1017/jie.2014.18 | photo-elicitation |
| Hinthorne, Lauren Leigh | A Picture Is Worth a Thousand Words: Using the Visual Interpretation Narrative Exercise to Elicit Non-Elite Perceptions of Democracy | 2012 | Field Methods | 24 | 3 | 348-364 | http://dx.doi.org/10.1177/1525822X12444065 | cartoons |
| Hodgetts, Darrin; Chamberlain, Kerry; Groot, Shiloh | Reflections on the visual in community research and action. | 2011 | Visual methods in psychology: Using and interpreting images in qualitative research. | | | 299-313 | | photo-elicitation |
| Holgate, Jane; Keles, Janroj; Kumarappan, Leena | Visualizing 'community': an experiment in participatory photography among Kurdish diasporic workers in London | 2012 | The Sociological Review | 60 | 2 | 312 | http://dx.doi.org/10.1111/j.1467-954X.2012.02075.x | photo-elicitation |
| Holland, Sierra | Pregnant with possibility: The importance of visual data in (re)presenting queer womenâ€™s experiences of reproduction | 2019 | Methodological Innovations | 12 | 1 | | http://dx.doi.org/10.1177/2059799119829429 | photo-elicitation |
| Holley, Jessica; Gillard, Steven | Developing and using vignettes to explore the relationship between risk management practice and recovery-oriented care in mental health services | 2018 | Qualitative Health Research | 28 | 3 | 371-380 | http://dx.doi.org/10.1177/1049732317725284 | vignette |
| Hollomotz Andrea | Successful interviews with people with intellectual disability | 2018 | Qualitative Research | 18 | 2 | 153-170 | http://dx.doi.org/10.1177/1468794117713810 | photo-elicitation; vignette |
| Holm, Gunilla | Photography as a research method. | 2014 | The Oxford handbook of qualitative research. | | | 380-402 | | photo-elicitation |
| Holt, Brenda Southerland | Creating Interpretive Visual Texts | 2012 | International Journal of Qualitative Studies in Education (QSE) | 25 | 5 | 665-679 | http://dx.doi.org/10.1080/09518398.2010.539580 | photo-elicitation |
| Impellizzeri, Jonathan; Savinsky, David M; King, John A; Leitch-Alford, Linda | Conceptual mapping task: An effective verification tool for qualitative counseling research. | 2017 | Counseling Outcome Research and Evaluation | 8 | 1 | 31-47 | http://dx.doi.org/10.1080/21501378.2017.1327745 | concept mapping |
| Jalan, Ishan | Researching dark emotions: Eliciting stories of envy. Methods of exploring emotions | 2015 | Routledge | | | 81-89 | | Review across categories |
| Janzen, Melanie D. | (Re)searching Methods: Reading Fiction in Literary Response Groups | 2015 | International Journal of Qualitative Studies in Education (QSE) | 28 | 8 | 989-1004 | http://dx.doi.org/10.1080/09518398.2014.940412 | storytelling |
| Jaspersen, Lena J; Stein, Christian | Beyond the matrix: Visual methods for qualitative network research. | 2019 | British Journal of Management | 30 | 3 | 748-763 | http://dx.doi.org/10.1111/1467-8551.12339 | mapping |
| Jehn, Karen A; Jonsen, Karsten | A multimethod approach to the study of sensitive organizational issues. | 2010 | Journal of Mixed Methods Research | 4 | 4 | 313-341 | http://dx.doi.org/10.1177/1558689810380920 | mapping |
| Jenkins, Nicholas; Bloor, Michael; Fischer, Jan; Berney, Lee; Neale, Joanne | Putting it in context: The use of vignettes in qualitative interviewing | 2010 | Qualitative Research | 10 | 2 | 175-198 | http://dx.doi.org/10.1177/1468794109356737 | vignette |
| Johnson, James E; Dong, Pool Ip | Methods of studying play. | 2019 | The Cambridge handbook of play: Developmental and disciplinary perspectives. | | | 399-415 | | photo-elicitation; drawing |
| Johnson, Katherine | Visualising mental health with an LGBT community group: Method, process, theory. | 2011 | Visual methods in psychology: Using and interpreting images in qualitative research. | | | 173-189 | | photo-elicitation |
| Johnson, Kayla M. | Hotdog as Metaphor: (Co)Developing Stories of Learning through Photo-Cued Interviewing | 2020 | Teachers College Record | 122 | 9 | | | photo-elicitation |
| Jones, Adam; Woolley, Janet | The email-diary: A promising research tool for the 21st century? | 2015 | Qualitative Research | 15 | 6 | 705-721 | http://dx.doi.org/10.1177/1468794114561347 | diary |
| Joseph, Tatiana | "We Have Rights, and We Can Make It Right:" Application of Photovoice as Tool of Empowerment with Low Income Latinx Students | 2017 | Journal of Ethnographic & Qualitative Research | 11 | 4 | 293-304 | | photo-elicitation; photovoice |
| Kalo, Zsuzsa; Racz, Jozsef | Map task, a new method for improving needle exchange services. | 2013 | Journal of Substance Use | 18 | 1 | 46-55 | http://dx.doi.org/10.3109/14659891.2012.735897 | mapping |

| | | | | | | | | |
|---|---|---|---|---|---|---|---|---|
| Kalter, Steve | Using projectives to uncover "Aha moments" in qualitative research. | 2016 | Qualitative research methods in consumer psychology: Ethnography and culture. | | | 131-146 | http://dx.doi.org/10.1207/S15327752JPA7903_03 | photo-elicitation |
| Kandemir, Asli; Budd, Richard | Using Vignettes to Explore Reality and Values With Young People | 2018 | Forum : Qualitative Social Research | 19 | 2 | | | vignette |
| Kang, Timothy; Kruttschnitt, Candace; Goodman, Philip | Multi-Method Synergy: Using the Life History Calendar and Life as a Film for Retrospective Narratives | 2017 | The Howard Journal of Crime and Justice | 56 | 4 | 532-553 | http://dx.doi.org/10.1111/hojo.12231 | life grid |
| Kaplan, Abram W. | Shifting Paradigms in Environmental Research Methods through the Visual Arts | 2020 | Arts and Humanities in Higher Education: An International Journal of Theory, Research and Practice | 19 | 2 | 115-143 | https://doi.org/10.1177/1474022218787158 | photo-elicitation |
| Kingsley, Joanne | Visual methodology in classroom inquiry: Enhancing complementary qualitative research designs. | 2009 | Alberta Journal of Educational Research | 55 | 4 | 534-548 | | photo-elicitation |
| Klein, Maike; Milner, Rebecca J | The use of body-mapping in interpretative phenomenological analyses: a methodological discussion | 2019 | International Journal of Social Research Methodology | 22 | 5 | 533-543 | http://dx.doi.org/10.1080/13645579.2019.1593378 | body mapping |
| Kolar Kat; Farah, Ahmad; Chan, Linda; Erickson, Patricia G | Timeline Mapping in Qualitative Interviews: A Study of Resilience with Marginalized Groups | 2015 | International Journal of Qualitative Methods | 14 | 3 | 13-32 | http://dx.doi.org/10.1177/160940691501400302 | timeline |
| Koopman-Boyden, Peggy; Richardson, Margaret | An evaluation of mixed methods (diaries and focus groups) when working with older people | 2013 | International Journal of Social Research Methodology: Theory & Practice | 16 | 5 | 389-401 | http://dx.doi.org/10.1080/13645579.2012.716971 | diary |
| Lahlou, Saadi; Le Bellu, Sophie; Boesen-Mariani, Sabine | Subjective Evidence Based Ethnography: Method and applications. | 2015 | Integrative Psychological & Behavioral Science | 49 | 2 | 216-238 | http://dx.doi.org/10.1007/s12124-014-9288-9 | video-elicitation |
| Lang, Michael; Laing, Catherine; Moules, Nancy; Estefan, Andrew | Words, Camera, Music, Action: A Methodology of Digital Storytelling in a Health Care Setting | 2019 | International Journal of Qualitative Methods | 18 | | | http://dx.doi.org/10.1177/1609406919863241 | story-telling |
| Lapatin, Sheri; Goncalves, Marta; Nillni, Anna; Chavez, Ligia; Quinn, Roxana Llerena; Green, Alexander; Alegria, Margarita | Lessons from the use of vignettes in the study of mental health service disparities. | 2012 | Health Services Research | 47 | 3, Pt2 | 1345-1362 | http://dx.doi.org/10.1111/j.1475-6773.2011.01360.x | vignette |
| Latz, Amanda O.; Phelps-Ward, Robin; Royer, Dan; Peters, Tiffany | Photovoice as Methodology, Pedagogy, and Partnership-Building Tool: A Graduate and Community College Student Collaboration | 2016 | Journal of Public Scholarship in Higher Education | 6 | | 124-142 | | photo-elicitation |
| Laukkanen, Mauri | Comparative Causal Mapping and CMAP3 Software in Qualitative Studies | 2012 | Forum Qualitative Sozialforschung/Forum: Qualitative Social Research | 13 | 2 | | | mapping |
| LeBaron, Curtis; Jarzabkowski, Paula; Pratt, Michael G; Fetzer, Greg | An introduction to video methods in organizational research. | 2018 | Organizational Research Methods | 21 | 2 | 239-260 | http://dx.doi.org/10.1177/1094428117745649 | video-elicitation |
| Lee-Ann, Fenge; Jones, Kip; Read, Rosie | Connecting Participatory Methods in a Study of Older Lesbian and Gay Citizens in Rural Areas | 2010 | International Journal of Qualitative Methods | 9 | 4 | 320-333 | http://dx.doi.org/10.1177/160940691000900402 | photo-elicitation |
| Leigh, Jadwiga; Morriss, Lisa; Morriss, Matthew | Making visible an invisible trade: Exploring the everyday experiences of doing social work and being a social worker | 2020 | Qualitative Social Work | 19 | 2 | 267-283 | http://dx.doi.org/10.1177/1473325018824629 | photo-elicitation |
| Levell Jade | â€œThose songs were the ones that made me, nobody asked me this question beforeâ€: Music Elicitation with ex-gang involved men about their experiences of childhood domestic violence and abuse | 2019 | International Journal of Qualitative Methods | 18 | | | http://dx.doi.org/10.1177/1609406919852010 | music videos |
| Li, Bing Yu; Ho Rainbow Tin Hung | Unveiling the Unspeakable: Integrating Video Elicitation Focus Group Interviews and Participatory Video in an Action Research Project on Dementia Care Development | 2019 | International Journal of Qualitative Methods | 18 | | | http://dx.doi.org/10.1177/1609406919830561 | video-elicitation |
| Lowery, Damon R; Morse, Wayde C | A Qualitative Method for Collecting Spatial Data on Important Places for Recreation, Livelihoods, and Ecological Meanings: Integrating Focus Groups with Public Participation Geographic Information Systems | 2013 | Society and Natural Resources | 26 | 12 | 1422-1437 | http://dx.doi.org/10.1080/08941920.2013.819954 | mapping |
| Lynch, Jonathan; Mannion, Greg | Enacting a Place-Responsive Research Methodology: Walking Interviews with Educators | 2016 | Journal of Adventure Education and Outdoor Learning | 16 | 4 | 330-345 | http://dx.doi.org/10.1080/14729679.2016.1163271 | walking |
| Majumdar, Anamika | Using photographs of places, spaces and objects to explore South Asian women's experience of close relationships and marriage. | 2011 | Visual methods in psychology: Using and interpreting images in qualitative research. | | | 69-84 | | photo-elicitation |
| Mannay, Dawn; Morgan, Melanie | Doing ethnography or applying a qualitative technique? Reflections from the 'waiting field'. | 2015 | Qualitative Research | 15 | 2 | 166-182 | http://dx.doi.org/10.1177/1468794113517391 | photo-elicitation |

| | | | | | | | | |
|---|---|---|---|---|---|---|---|---|
| Mapp, Fiona | Reflections on Using a Flash Card Activity for Studying Social Representations of Sexually Transmitted Infections | 2017 | International Journal of Qualitative Methods | 16 | 1 | | http://dx.doi.org/10.1177/1609406917703741 | drawing (flash cards) |
| Marsh, Wendy; Leamon, Jen; Robinson, Ann; Shawe, Jill | LEARNS: A creative approach to analysing and representing narrative data incorporating photo-elicitation techniques. | 2018 | Journal of Research in Nursing | 23 | 4 | 334-343 | http://dx.doi.org/10.1177/1744987117750218 | photo-elicitation |
| Martiniello, Marco | Visual sociology approaches in migration, ethnic and racial studies | 2017 | Ethnic and Racial Studies | 40 | 8 | 1184-1190 | http://dx.doi.org/10.1080/01419870.2017.1295163 | Review across categories |
| McDonnell, Liz; Scott, Susie; Dawson, Matt | A multidimensional view? Evaluating the different and combined contributions of diaries and interviews in an exploration of asexual identities and intimacies | 2017 | Qualitative Research | 17 | 5 | 520-536 | http://dx.doi.org/10.1177/1468794116676516 | diary |
| McGrath, Laura; Mullarkey, Shauna; Reavey, Paula | Building visual worlds: Using maps in qualitative psychological research on affect and emotion. | 2020 | Qualitative Research in Psychology | 17 | 1 | 75-97 | http://dx.doi.org/10.1080/14780887.2019.1577517 | mapping |
| Meck, Chongo; Chase, Robert M; Lavoie, JosÃ©e G; Harder, H G; Mignone, Javier | The Life Story Board as a Tool for Qualitative Research | 2018 | International Journal of Qualitative Methods | 17 | 1 | | http://dx.doi.org/10.1177/1609406917752440 | life grid |
| Melnikova, Olga T; Khoroshilov, Dmitry A | Priority research directions in the area of qualitative methodology. | 2010 | Psychology in Russia: State of the Art | 3 | | 46-72 | http://dx.doi.org/10.11621/pir.2010.0002 | drawing |
| Metcalfe, Amy Scott | Visual Juxtaposition as Qualitative Inquiry in Educational Research | 2015 | International Journal of Qualitative Studies in Education (QSE) | 28 | 2 | 151-167 | http://dx.doi.org/10.1080/09518398.2013.855340 | photo-elicitation |
| Migliorini, Laura; Rania, Nadia | A qualitative method to "make visible" the world of intercultural relationships: The photovoice in social psychology. | 2017 | Qualitative Research in Psychology | 14 | 2 | 131-145 | http://dx.doi.org/10.1080/14780887.2016.1263698 | photo-elicitation |
| Mignone, Javier; Chase, Robert M; Roger, Kerstin | A Graphic and Tactile Data Elicitation Tool for Qualitative Research: The Life Story Board | 2019 | Forum : Qualitative Social Research | 20 | 2 | | | life grid |
| Mitchell, Claudia; Lamb, Pamela; Raissadat Haleh | Exploring the Impact of Youth-Produced Images on Family, Community, and Policy | 2018 | International Journal of Qualitative Methods | 17 | 1 | | http://dx.doi.org/10.1177/1609406918807609 | vignette; photo-elicitation |
| Mitchell, Michael A; Hedayati, Daniel O; Rodriguez, Keri L; Gordon, Adam J; Broyles, Lauren M; True, Gala; Balbale, Salva N; Conley, James W | Logistical Lessons Learned in Designing and Executing a Photo-Elicitation Study in the Veterans Health Administration | 2016 | The Qualitative Report | 21 | 7 | 1303-1315 | | photo-elicitation |
| Molden, Olivia C. | Short Take: Story-Mapping Experiences | 2020 | Field Methods | 32 | 2 | 131-139 | https://doi.org/10.1177/1525822X19877381 | story-telling; mapping |
| Mulvale Gillian; Moll, Sandra; Miatello Ashleigh; Murray-Leung, Louise; Rogerson Karlie; Sassi, Roberto B | Co-designing Services for Youth With Mental Health Issues: Novel Elicitation Approaches | 2019 | International Journal of Qualitative Methods | 18 | | | http://dx.doi.org/10.1177/1609406918816244 | mapping |
| Mulvey, Michael S; Kavalam, Beena E | Mining deeper meaning in consumer decision maps. | 2010 | Qualitative Market Research: An International Journal | 13 | 4 | 372-388 | http://dx.doi.org/10.1108/13522751011078809 | metaphor |
| Murawska, Jaclyn M.; Walker, David A. | Visual Tools for Eliciting Connections and Cohesiveness in Mixed Methods Research | 2017 | Mid-Western Educational Researcher | 29 | 3 | 274-290 | | diagram |
| Murray, Linda; Nash, Meredith | The challenges of participant photography: A critical reflection on methodology and ethics in two cultural contexts. | 2017 | Qualitative Health Research | 27 | 6 | 923-937 | http://dx.doi.org/10.1177/1049732316668819 | photo-elicitation |
| Nathan Stephens Griffin | Comics and visual biography: sequential art in social research | 2019 | Visual Studies | 34 | 4 | 319-335 | http://dx.doi.org/10.1080/1472586X.2019.1691940 | comic |
| Nicholas, Maria | Affordances of Using Multiple Videoed Events to Construct a Rich Understanding of Adult-Child Book Readings | 2018 | International Journal of Research & Method in Education | 41 | 2 | 125-141 | https://doi.org/10.1080/1743727X.2016.1254176 | video-elicitation |
| Nind, Melanie; Kilburn, Daniel; Wiles, Rose | Using Video and Dialogue to Generate Pedagogic Knowledge: Teachers, Learners and Researchers Reflecting Together on the Pedagogy of Social Research Methods | 2015 | International Journal of Social Research Methodology | 18 | 5 | 561-576 | https://doi.org/10.1080/13645579.2015.1062628 | video-elicitation; stimulated recall |
| Nind, Melanie; Vinha, Hilra | Creative interactions with data: using visual and metaphorical devices in repeated focus groups | 2016 | Qualitative Research | 16 | 1 | 26-Sep | http://dx.doi.org/10.1177/1468794114557993 | poetry; metaphor |
| Nordstrom, Susan Naomi | Object-Interviews: Folding, Unfolding, and Refolding Perceptions of Objects | 2013 | International Journal of Qualitative Methods | 12 | 1 | 237-257 | http://dx.doi.org/10.1177/160940691301200111 | object-elicitation |
| Novek, Sheila; Morris-Oswald, Toni; Menec, Verena | Using photovoice with older adults: Some methodological strengths and issues. | 2012 | Ageing & Society | 32 | 3 | 451-470 | http://dx.doi.org/10.1017/S0144686X11000377 | photo-elicitation |
| Nykiforuk Candace IJ; Vallianatos, Helen; Nieuwendyk, Laura M | Photovoice as a Method for Revealing Community Perceptions of the Built and Social Environment | 2011 | International Journal of Qualitative Methods | 10 | 2 | 103-124 | http://dx.doi.org/10.1177/160940691101000201 | photo-elicitation |
| O'Brien, Mark; Varga-Atkins, Tunde; Umoquit, Muriah; Tso, Peggy | Cultural-Historical Activity Theory and "The Visual" in Research: Exploring the Ontological Consequences of the Use of Visual Methods | 2012 | International Journal of Research & Method in Education | 35 | 3 | 251-268 | http://dx.doi.org/10.1080/1743727X.2012.717433 | drawing |

| | | | | | | | | |
|---|---|---|---|---|---|---|---|---|
| O'Malley, Lori J.; Munsell, Sonya E. | PhotoVoice: An Innovative Qualitative Method in Research and Classroom Teaching | 2020 | Educational Research: Theory and Practice | 31 | 1 | 26-32 | | photo-elicitation; photovoice |
| Orr, Noreen; Phoenix, Cassandra | Photographing physical activity: using visual methods to 'grasp at' the sensual experiences of the ageing body | 2015 | Qualitative Research | 15 | 4 | 454-472 | http://dx.doi.org/10.1177/1468794114543401 | photo-elicitation |
| Ortega-Alcazar, Iliana; Dyck, Isabel | Migrant narratives of health and well-being: Challenging 'othering' processes through photo-elicitation interviews. | 2012 | Critical Social Policy | 32 | 1 | 106-125 | http://dx.doi.org/10.1177/0261018311425981 | photo-elicitation |
| Osei-Kofi, Nana | The Emancipatory Potential of Arts-Based Research for Social Justice | 2013 | Equity & Excellence in Education | 46 | 1 | 135-149 | http://dx.doi.org/10.1080/10665684.2013.750202 | photo-elicitation |
| Ostby, May; Bjorkly, Stal | Vignette selection for ethical reflections: A selection procedure for vignettes to investigate staff reflections on the ethical challenges in interaction with people with intellectual disabilities. | 2011 | Ethics and Social Welfare | 5 | 3 | 277-295 | http://dx.doi.org/10.1080/17496535.2010.550129 | vignette |
| O'Toole, Paddy | Capturing Undergraduate Experience through Participant-Generated Video | 2013 | The Qualitative Report | 18 | 33 | | | diary |
| Ottmann, Goetz; Crosbie, Jenny | Mixed Method Approaches in Open-Ended, Qualitative, Exploratory Research Involving People with Intellectual Disabilities: A Comparative Methods Study | 2013 | Journal of Intellectual Disabilities | 17 | 3 | 182-197 | http://dx.doi.org/10.1177/1744629513494927 | photo-elicitation |
| Owens, Christabel; Carter, Mary; Shenton, Deborah; Byng, Richard; Quinn, Cath | Engaging without exposing: Use of a fictional character to facilitate mental health talk in focus groups with men who have been subject to the criminal justice system | 2018 | Qualitative Health Research | 28 | 13 | 2102-2114 | http://dx.doi.org/10.1177/1049732318785359 | vignette |
| Owens, Otis L; Beer, Jenay M; Revels Asa; Levkoff, Sue | Feasibility of using a video diary methodology with older African Americans living alone | 2019 | Qualitative Social Work | 18 | 3 | 397-416 | http://dx.doi.org/10.1177/1473325017729570 | diary |
| Ozanne, Julie L; Moscato, Emily M; Kunkel, Danylle R | Transformative photography: Evaluation and best practices for eliciting social and policy changes. | 2013 | Journal of Public Policy & Marketing | 32 | 1 | 45-65 | http://dx.doi.org/10.1509/jppm.11.161 | photo-elicitation |
| Pain, Helen | A Literature Review to Evaluate the Choice and Use of Visual Methods | 2012 | International Journal of Qualitative Methods | 11 | 4 | 303-319 | http://dx.doi.org/10.1177/160940691201100401 | Review across categories |
| Palmer, Victoria J; Dowrick, Christopher; Gunn, Jane M | Mandalas as a visual research method for understanding primary care for depression | 2014 | International Journal of Social Research Methodology | 17 | 5 | 527-541 | http://dx.doi.org/10.1080/13645579.2013.796764 | drawing (graphic-elicitation) |
| Papaloukas, Periklis; Quincey, Kerry; Williamson, Iain R | Venturing into the visual voice: Combining photos and interviews in phenomenological inquiry around marginalisation and chronic illness. | 2017 | Qualitative Research in Psychology | 14 | 4 | 415-441 | http://dx.doi.org/10.1080/14780887.2017.1329364 | photo-elicitation |
| Paton, Joy; Horsfall, Debbie; Carrington Amie | Sensitive Inquiry in Mental Health | 2018 | International Journal of Qualitative Methods | 17 | 1 | | http://dx.doi.org/10.1177/1609406918761422 | photo-elicitation |
| Pauwels, Luc | Visual Sociology Reframed: An Analytical Synthesis and Discussion of Visual Methods in Social and Cultural Research | 2010 | Sociological Methods & Research | 38 | 4 | 545-581 | http://dx.doi.org/10.1177/0049124110366233 | Review across categories |
| Pearce, Susie; Gibson, Faith; Whelan, Jeremy; Kelly, Daniel | Untellable tales and uncertain futures: the unfolding narratives of young adults with cancer | 2020 | International Journal of Social Research Methodology | 23 | 4 | 377-390 | http://dx.doi.org/10.1080/13645579.2020.1719614 | photo-elicitation |
| Pettinger Clare; Letherby Gayle; Parsons, Julie M; Withers Lyndsey; Cunningham, Miranda; Whiteford, Andrew; Dâ€™Aprano Gia; Ayres, Richard; Sutton, Carole | Employing participatory methods to engage an under-researched group: Opportunities and challenges | 2018 | Methodological Innovations | 11 | 1 | | http://dx.doi.org/10.1177/2059799118769820 | photo-elicitation |
| Phillipson Lyn; Hammond, Athena | More Than Talking | 2018 | International Journal of Qualitative Methods | 17 | 1 | | http://dx.doi.org/10.1177/1609406918782784 | photo-elicitation |
| Phoenix, Ann; Brannen, Julia | Researching family practices in everyday life: methodological reflections from two studies | 2014 | International Journal of Social Research Methodology | 17 | 1 | | http://dx.doi.org/10.1080/13645579.2014.854001 | photo-elicitation |
| Pilcher, Katy; Martin, Wendy; Williams, Veronika | Issues of Collaboration, Representation, Meaning and Emotions: Utilising Participant-Led Visual Diaries to Capture the Everyday Lives of People in Mid to Later Life | 2016 | International Journal of Social Research Methodology | 19 | 6 | 677-692 | http://dx.doi.org/10.1080/13645579.2015.1086199 | photo-elicitation |
| Pink, Sarah | Multimodality, multisensoriality and ethnographic knowing: social semiotics and the phenomenology of perception | 2011 | Qualitative Research | 11 | 3 | 261-276 | http://dx.doi.org/10.1177/1468794111399835 | images |
| Pithouse-Morgan, Kathleen; Naicker, Inbanathan; Chikoko, Vitallis; Pillay, Daisy; Morojele, Pholoho; Hlao, Teboho | Entering an Ambiguous Space: Evoking Polyvocality in Educational Research through Collective Poetic Inquiry | 2014 | Perspectives in Education | 32 | 4 | 149-170 | | poetry |
| Pitt, Edd | Drawing: A visual method as an expressive data collection technique. | 2017 | Teaching visual methods in the social sciences. | | | 84-100 | | drawing |
| Polisenska, Veronika A | Interviewing offenders in a penitentiary environment and the use of mental maps during interviews. | 2010 | Offenders on offending: Learning about crime from criminals. | | | 273-289 | | mapping |
| Powell, Kimberly | Making Sense of Place: Mapping as a Multisensory Research Method | 2010 | Qualitative Inquiry | 16 | 7 | 539-555 | http://dx.doi.org/10.1177/1077800410372600 | mapping |
| Pristed Nielsen, Helene; MÃ¸ller, Karina Torp | Studying place practices and consumption through volunteer-employed photography | 2016 | Journal of Consumer Culture | 16 | 3 | 781-800 | http://dx.doi.org/10.1177/1469540514536195 | photo-elicitation |

| | | | | | | | | |
|---|---|---|---|---|---|---|---|---|
| Provenzo, Eugene F.; Ameen, Edward; Bengochea, Alain; Doorn, Kristen; Pontier, Ryan; Sembiante, Sabrina | Photography and Oral History as a Means of Chronicling the Homeless in Miami: The "StreetWays" Project | 2011 | Educational Studies: Journal of the American Educational Studies Association | 47 | 5 | 419-435 | | photo-elicitation |
| Qui√±ones, Gloria; Ridgway, Avis; Li, Liang | Collaborative Drawing: A Creative Tool for Examination of Infant-Toddler Pedagogical Practices | 2019 | Australasian Journal of Early Childhood | 44 | 3 | 230-243 | https://doi.org/10.1177/1836939119855219 | drawing |
| Raby, Rebecca; Lehmann, Wolfgang; Helleiner, Jane; Easterbrook Riley | Reflections on Using Participant-Generated, Digital Photo-Elicitation in Research With Young Canadians About Their First Part-Time Jobs | 2018 | International Journal of Qualitative Methods | 17 | 1 | | http://dx.doi.org/10.1177/1609406918790681 | photo-elicitation |
| Rainford, Jon | Confidence and the effectiveness of creative methods in qualitative interviews with adults | 2020 | International Journal of Social Research Methodology | 23 | 1 | 109-122 | http://dx.doi.org/10.1080/13645579.2019.1672287 | drawing |
| Ramvi, Ellen; Manley, Julian; Froggett, Lynn; Liveng, Anne; Lading, Aase; Hollway, Wendy; Gripsrud, Birgitta Haga | The visual matrix method in a study of death and dying: Methodological reflections. | 2019 | Psychoanalysis, Culture & Society | 24 | 1 | 31-52 | http://dx.doi.org/10.1057/s41282-018-0095-y | images |
| Ravn, Signe; Duff, Cameron | Putting the party down on paper: A novel method for mapping youth drug use in private settings. | 2015 | Health & Place | 31 | | 124-132 | http://dx.doi.org/10.1016/j.healthplace.2014.11.010 | mapping |
| Reid, David A; Simmt, Elaine; Savard, Annie; Suurtamm, Christine; Manuel, Dominic; Lin, Terry Wan Jung; Quigley, Brenna; Knipping, Christine | Observing Observers: Using Video to Prompt and Record Reflections on Teachers' Pedagogies in Four Regions of Canada | 2015 | Research in Comparative and International Education | 10 | 3 | 367-382 | https://doi.org/10.1177/1745499915580425 | video-elicitation |
| Richardson, Tobin; Latz, Amanda O.; Coren, Ashleigh; Pickens, Chanelle | Mapping Success: Understanding the Early Educational Experiences of Medical Students through the Creation of Chronological Educational Maps | 2016 | Journal of Ethnographic & Qualitative Research | 11 | 2 | 136-153 | | mapping |
| Rizvi, Sana | Using Fiction to Reveal Truth: Challenges of Using Vignettes to Understand Participant Experiences Within Qualitative Research | 2019 | Forum : Qualitative Social Research | 20 | 1 | | | vignette |
| Rodriguez, Sheri K; Kerrigan, Monica Reid | Using Graphic Elicitation to Explore Community College Transfer Student Identity, Development, and Engagement | 2016 | The Qualitative Report | 21 | 6 | 1052-1070 | | mapping |
| Roos, Vera | Implementing the Mmogo-method: A group of Setswana-speaking older people's relational experiences in a rural community setting. | 2016 | Understanding relational and group experiences through the Mmogo-method. | | | 55-88 | http://dx.doi.org/10.1007/978-3-319-31224-8_5 | crafting; object-making |
| Rosenblum, Jason; Hughes, Joan E. | Digital Recording Technologies in Phenomenological Investigations | 2017 | Journal of Ethnographic & Qualitative Research | 12 | 1 | 29-49 | | video-elicitation |
| Roth, Wendy D | Studying ethnic schemas: Integrating cognitive schemas into ethnicity research through photo elicitation. | 2015 | Studying ethnic identity: Methodological and conceptual approaches across disciplines. | | | 89-118 | http://dx.doi.org/10.1037/14618-005 | photo-elicitation |
| Roth, Wendy D. | Studying ethnic schemas: integrating cognitive schemas into ethnicity research through photo elicitation | 2015 | | | | | | photo-elicitation |
| Rowley, Jennifer; Jones, Rosalind; Vassiliou, Magda; Hanna, Sonya | Using card-based games to enhance the value of semi-structured interviews. | 2012 | International Journal of Market Research | 54 | 1 | 93-110 | http://dx.doi.org/10.2501/IJMR-54-1-093-110 | card task |
| Rumpf, CesraÃ©a | Decentering Power in Research with Criminalized Women: A Case for Photo-Elicitation Interviewing | 2017 | Sociological Focus | 50 | 1 | 18-35 | http://dx.doi.org/10.1080/00380237.2016.1218214 | photo-elicitation |
| SÃ¶derstrÃ¶m, Johanna | Life diagrams: a methodological and analytical tool for accessing life histories | 2020 | Qualitative Research | 20 | 1 | | http://dx.doi.org/10.1177/1468794118819068 | life grid |
| Sadler, Kirsten; Selkrig, Mark; Manathunga, Catherine | Teaching Is ... Opening up Spaces to Explore Academic Work in Fluid and Volatile Times | 2017 | Higher Education Research and Development | 36 | 1 | 171-186 | http://dx.doi.org/10.1080/07294360.2016.1171299 | postcards |
| Schiffer, Eva; Hauck, Jennifer | Net-Map: Collecting social network data and facilitating network learning through participatory influence network mapping. | 2010 | Field Methods | 22 | 3 | 231-249 | http://dx.doi.org/10.1177/1525822X10374798 | mapping |
| Schulze, SalomÃ© | The Value of Two Modes of Graphic Elicitation Interviews to Explore Factors That Impact on Student Learning in Higher Education | 2017 | Qualitative Sociology Review | 13 | 2 | | | mapping |
| Shaw, Donna | A New Look at an Old Research Method: Photo-Elicitation | 2013 | TESOL Journal | 4 | 4 | 785-799 | http://dx.doi.org/10.1002/tesj.108 | photo-elicitation |
| Shell, Lynn | Photo-Elicitation with Autodriving in Research with Individuals with Mild to Moderate Alzheimer's Disease: Advantages and Challenges | 2014 | International Journal of Qualitative Methods | 13 | 1 | 170-184 | http://dx.doi.org/10.1177/160940691401300106 | photo-elicitation |
| Sheridan, Joanna; Chamberlain, Kerry | The power of things. | 2011 | Qualitative Research in Psychology | 8 | 4 | 315-332 | http://dx.doi.org/10.1080/14780880903490821 | photo-elicitation; diary |
| Sheridan, Joanna; Chamberlain, Kerry; Dupuis, Ann | Timelining: visualizing experience | 2011 | Qualitative Research | 11 | 5 | 552-569 | http://dx.doi.org/10.1177/1468794111413235 | timeline |
| Shirani, Fiona; Parkhill, Karen; Butler, Catherine; Groves, Chris; Pidgeon, Nick; Henwood, Karen | Asking about the Future: Methodological Insights from Energy Biographies | 2016 | International Journal of Social Research Methodology | 19 | 4 | 429-444 | http://dx.doi.org/10.1080/13645579.2015.1029208 | photo-elicitation |
| Sidorenko Ewa | â€˜A Portrait of Lower Silesiaâ€™: Researching identity through collodion photography and memory narratives | 2019 | Methodological Innovations | 12 | 3 | | http://dx.doi.org/10.1177/2059799119890787 | photo-elicitation |

| | | | | | | | | |
|---|---|---|---|---|---|---|---|---|
| Sievers, Burkard | Thinking organisations through photographs: The social photo-matrix as a method for understanding organisations in depth. | 2013 | Socioanalytic methods: Discovering the hidden in organisations and social systems. | | | 129-151 | | photo-elicitation |
| Sievers, Burkard | It is difficult to think in the slammer: A social photo-matrix in a penal institution. | 2014 | The psychosocial and organization studies: Affect at work. | | | 129-157 | http://dx.doi.org/10.1057/9781137347855.0012 | photo-elicitation |
| Slee, Phillip; Skrzypiec, Grace; Sandhu, Damanjit; Kaur, Kirandeep; Campbell, Marilyn | PhotoStory: A legitimate research tool in cross-cultural research. | 2018 | Bullying, cyberbullying and student well-being in schools: Comparing European, Australian and Indian perspectives. | | | 189-207 | http://dx.doi.org/10.1017/9781316987384.012 | photo-elicitation |
| Snyder, Catherine | A Case Study of a Case Study: Analysis of a Robust Qualitative Research Methodology | 2012 | Qualitative Report | 17 | | | | photo-elicitation |
| Sopcak, Nicolette; Mayan, Maria; Skrypnek, Berna J | Engaging Young Fathers in Research through Photo-Interviewing | 2015 | The Qualitative Report | 20 | 11 | 1871-1880 | | photo-elicitation |
| Spokes, Matthew; Denham, Jack | Developing Interactive Elicitation: Social Desirability Bias and Capturing Play | 2019 | The Qualitative Report | 24 | 4 | 781-794 | | photo-elicitation; vignette |
| Spowart, Lucy; Nairn, Karen | (Re)performing emotions in diary-interviews | 2014 | Qualitative Research | 14 | 3 | 327-340 | http://dx.doi.org/10.1177/1468794112473498 | diary |
| Stevenson, Blair | Third spaces and video-stimulated recall: An exploration of teachersâ €™ cultural role in an Indigenous education context | 2015 | Educational Action Research | 23 | 2 | 290-305 | http://dx.doi.org/10.1080/09650792.2014.990986 | stimulated recall |
| Stieber, Roger Kerstin; Blomgren, Constance | Elicitation as a Mind-Set: Why Visual Data Matter? | 2019 | International Journal of Qualitative Methods | 18 | | | http://dx.doi.org/10.1177/1609406919835378 | life grid; photo-elicitation |
| Stravakou, Pelagia A; Lozgka, Evangelia Ch | Vignettes in Qualitative Educational Research: Investigating Greek School Principals' Values | 2018 | The Qualitative Report | 23 | 5 | 1188-1207 | | vignette |
| Sum, Raymond Kim Wai; Shi, Teng-Yao | Lived Experiences of a Hong Kong Physical Education Teacher: Ethnographical and Phenomenological Approaches | 2016 | The Qualitative Report | 21 | 1 | 127-142 | | photo-elicitation |
| Sutton, Barbara | Playful cards, serious talk: A qualitative research technique to elicit women's embodied experiences. | 2011 | Qualitative Research | 11 | 2 | 177-196 | http://dx.doi.org/10.1177/1468794110394070 | card task |
| Switzer, S; Guta, A; de Prinse, K; Carusone, S Chan; Strike, C | Visualizing harm reduction: Methodological and ethical considerations | 2015 | Social Science & Medicine | 133 | | 77 | http://dx.doi.org/10.1016/j.socscimed.2015.03.040 | photo-elicitation |
| Switzer, Sarah | Working With Photo Installation and Metaphor: Re-Visioning Photovoice Research | 2019 | International Journal of Qualitative Methods | 18 | | | http://dx.doi.org/10.1177/1609406919872395 | photo-elicitation |
| Tarr, Jen; Cornish, Flora; Gonzalez-Polledo, Elena | Beyond the binaries: Reshaping pain communication through arts workshops. | 2018 | Sociology of Health & Illness | 40 | 3 | 577-592 | http://dx.doi.org/10.1111/1467-9566.12669 | drawing; photo-elicitation |
| Tarr, Jen; Thomas, Helen | Mapping embodiment: methodologies for representing pain and injury | 2011 | Qualitative Research | 11 | 2 | 141-157 | http://dx.doi.org/10.1177/1468794110394067 | body mapping |
| Teti, Michelle; Murray, Cynthia; Johnson, LaShaune; Binson, Diane | Photovoice as a community-based participatory research method among women living with HIV/AIDS: Ethical opportunities and challenges. | 2012 | Journal of Empirical Research on Human Research Ethics | 7 | 4 | 34-43 | http://dx.doi.org/10.1525/jer.2012.7.4.34 | photo-elicitation |
| Thompson, MaryEllen; Oelker Abigail | Use of Participant-Generated Photographs versus Time Use Diaries as a Method of Qualitative Data Collection | 2013 | International Journal of Qualitative Methods | 12 | 1 | 624-637 | http://dx.doi.org/10.1177/160940691301200133 | diary; photo-elicitation |
| Thygesen, Marianne K; Pedersen, Birthe D; Kragstrup, Jakob; Wagner, Lis; Mogensen, Ole | Utilizing a New Graphical Elicitation Technique to Collect Emotional Narratives Describing Disease Trajectories | 2011 | The Qualitative Report | 16 | 2 | 596-608 | | timeline |
| Tishelman, Carol; Lindqvist, Olav; Hajdarevic, Senada; Rasmussen, Birgit H; Goliath, Ida | Beyond the visual and verbal: Using participant-produced photographs in research on the surroundings for care at the end-of-life | 2016 | Social Science & Medicine | 168 | | 120 | http://dx.doi.org/10.1016/j.socscimed.2016.09.012 | photo-elicitation |
| Torre, Daniela; Murphy, Joseph | A Different Lens: Using Photo-Elicitation Interviews in Education Research | 2015 | Education Policy Analysis Archives | 23 | 111 | | | photo-elicitation |
| Tour, Ekaterina | Understanding Digital Literacy Practices: What Can Be Learnt with Visual Methods? | 2017 | Changing English: Studies in Culture and Education | 24 | 4 | 413-425 | http://dx.doi.org/10.1080/1358684X.2017.1304815 | photo-elicitation |
| Turk, Melanie T; Fapohunda, Abimbola; Zoucha, Rick | Using Photovoice to explore Nigerian immigrants' eating and physical activity in the United States. | 2015 | Journal of Nursing Scholarship | 47 | 1 | 16-24 | http://dx.doi.org/10.1111/jnu.12105 | photo-elicitation |
| Umoquit, Muriah; Tso, Peggy; Varga-Atkins, TÃ¼nde; O'Brien, Mark; Wheeldon, Johannes | Diagrammatic Elicitation: Defining the Use of Diagrams in Data Collection | 2013 | The Qualitative Report | 18 | 30 | | | drawing |
| van Schalkwyk, Gertina J. | Collage Life Story Elicitation Technique: A Representational Technique for Scaffolding Autobiographical Memories | 2010 | Qualitative Report | 15 | 3 | 675-695 | | collage; life grid |
| Vesterinen, Olli; Toom, Auli; Patrikainen, Sanna | The Stimulated Recall Method and ICTs in Research on the Reasoning of Teachers | 2010 | International Journal of Research & Method in Education | 33 | 2 | 183-197 | | stimulated recall |

| | | | | | | | | |
|---|---|---|---|---|---|---|---|---|
| Vigurs, Katy; Kara, Helen | Participants' Productive Disruption of a Community Photo-Elicitation Project: Improvised Methodologies in Practice | 2017 | International Journal of Social Research Methodology | 20 | 5 | 513-523 | http://dx.doi.org/10.1080/13645579.2016.1221259 | photo-elicitation |
| VilÃ , Montserrat; Pallisera, Maria; Fullana, Judit | Exploring the Present and Projecting the Future: People with Severe Mental Illness Speaking for Themselves | 2016 | International Journal of Qualitative Studies in Education (QSE) | 29 | 9 | 1118-1130 | http://dx.doi.org/10.1080/09518398.2016.1201164 | photo-elicitation |
| Wallace, Heather Julie; McDonald, Susan; Belton, Suzanne; Miranda, Agueda Isolina; da Costa, Eurico; da Conceicao Matos, Livio; Henderson, Helen; Taft, Angela | Body mapping to explore reproductive ethno-physiological beliefs and knowledge of contraception in Timor-Leste. | 2018 | Qualitative Health Research | 28 | 7 | 1171-1184 | http://dx.doi.org/10.1177/1049732317750382 | body mapping |
| Warner, Elyse; Johnson, Louise; Andrews, Fiona | Exploring the Suburban Ideal | 2016 | International Journal of Qualitative Methods | 15 | 1 | | http://dx.doi.org/10.1177/1609406916654716 | photo-elicitation |
| Wass, Rob; Anderson, Vivienne; Rabello, Rafaela; Golding, Clinton; Rangi, Ana; Eteuati, Esmay | Photovoice as a Research Method for Higher Education Research | 2020 | Higher Education Research and Development | 39 | 4 | 834-850 | https://doi.org/10.1080/07294360.2019.1692791 | photo-elicitation; photovoice |
| Wheeldon, J | Mapping mixed methods research: Methods, measures, and meaning. | 2010 | Journal of Mixed Methods Research | 4 | 2 | 87-102 | http://dx.doi.org/10.1177/1558689809358755 | concept mapping; mind mapping |
| Wheeldon, Johannes | Is a Picture Worth a Thousand Words? Using Mind Maps to Facilitate Participant Recall in Qualitative Research | 2011 | Qualitative Report | 16 | 2 | 509-522 | | concept mapping; mind mapping |
| White, Darcy; Stephenson, Rob | Using community mapping to understand family planning behavior. | 2014 | Field Methods | 26 | 4 | 406-420 | http://dx.doi.org/10.1177/1525822X14529256 | mapping |
| Whitworth, Andrew; Torras I Calvo, Maria Carme; Moss, Bodil; Amlesom Kifle, Nazareth; BlÃ¥sternes, Terje | Changing Libraries: Facilitating Self-Reflection and Action Research on Organizational Change in Academic Libraries | 2014 | New Review of Academic Librarianship | 20 | 2 | 251-274 | http://dx.doi.org/10.1080/13614533.2014.912989 | mapping |
| Wilhoit, Elizabeth D | Photo and video methods in organizational and managerial communication research. | 2017 | Management Communication Quarterly | 31 | 3 | 447-466 | http://dx.doi.org/10.1177/0893318917704511 | photo-elicitation |
| Wilhoit, Elizabeth D; Kisselburgh, Lorraine G | Through the eyes of the participant: Making connections between researcher and subject with participant viewpoint ethnography. | 2016 | Field Methods | 28 | 2 | 208-226 | http://dx.doi.org/10.1177/1525822X15601950 | video-elicitation |
| Williams Carawan, Lena; Nalavany, Blace | Using Photography and Art in Concept Mapping Research with Adults with Dyslexia | 2010 | Disability & Society | 25 | 3 | 317-329 | | concept mapping; photo-elicitation; collage |
| Williams, Roman R; Whitehouse, Kyle | Photo Elicitation and the Visual Sociology of Religion | 2015 | Review of Religious Research | 57 | 2 | 303-318 | http://dx.doi.org/10.1007/s13644-014-0199-5 | photo-elicitation |
| Williams, Sion; Keady, John | Centre Stage Diagramming: Late-stage Parkinson's disease and Alzheimer's disease | 2012 | Journal of Aging Studies | 26 | 2 | 204-213 | http://dx.doi.org/10.1016/j.jaging.2011.12.005 | diagram |
| Willig, Carla | Reflections on the use of object elicitation. | 2017 | Qualitative Psychology | 4 | 3 | 211-222 | http://dx.doi.org/10.1037/qup0000054 | object-elicitation |
| Wills, Wendy J; Dickinson, Angela M; Meah, Angela; Short, Frances | Reflections on the use of visual methods in a qualitative study of domestic kitchen practices. | 2016 | Sociology | 50 | 3 | 470-485 | http://dx.doi.org/10.1177/0038038515587651 | photo-elicitation; diary; collage |
| Zundel, Mike; MacIntosh, Robert; Mackay, David | The utility of video diaries for organizational research | 2018 | Organizational Research Methods | 21 | 2 | 386-411 | http://dx.doi.org/10.1177/1094428116665463 | diary |
