## Appendix D for "More Than Words: An Integrative Review of Innovative Elicitation Techniques for Qualitative Interviews"

**How-to Guide Citations for Elicitation Techniques**

| **Technique** | **How-to Guide** |
|---|---|
| Concept mapping | Umoquit MJ, Tso P, Tünde VA, O'brien M, Wheeldon J. Diagrammatic elicitation: Defining the use of diagrams in data collection. The Qualitative Report. 2013;18(60):1-12. |
| Crafting | N/A |
| Drawing | Cristancho S, Bidinosti S, Lingard L, Novick R, Ott M, Forbes T. Seeing in different ways: Introducing "rich pictures"in the study of expert judgment. *Qualitative Health Research*. 2015;25(5):713-725. doi:10.1177/1049732314553594<br><br>Cristancho S, Helmich E. Rich pictures: a companion method for qualitative research in medical education. *Medical Education*. 2019;53(9):916-924. doi:10.1111/medu.13890 |
| Music-elicitation | N/A |
| Object-elicitation | Willig, C. Reflections on the use of object elicitation. Qualitative Psychology. 2017;4(3), 211–22. a different Willig one, but maybe more description/reflection on object elicitation itself. |
| Photo and video-elicitation | Barry, W., & Beighton, C., (2021). Using photovoice in participatory educational research. In Sage Research Methods Cases Part 1. SAGE Publications, Ltd., https://doi.org/10.4135/9781529758337<br><br>Boucher Jr ML, editor. Participant empowerment through photo-elicitation in ethnographic education research: New perspectives and approaches. Springer; 2017 Oct 13.<br><br>Camille A. Sutton-Brown (2014) Photovoice: A Methodological Guide, Photography and Culture, 7:2, 169-185, DOI: 10.2752/175145214X13999922103165<br><br>Cristancho S, LaDonna K, Field E. Visual methods in health professions research: purpose, challenges and opportunities. In: Cleland J, Durning SJ, eds. *Researching Medical Education*. 1st ed. Wiley; 2022:139-151. doi:10.1002/9781119839446.ch13<br><br>Zhang, Y., & Hennebry-Leung, M. (2023). A Review of Using Photo-Elicitation Interviews in Qualitative Education Research. International Journal of Qualitative Methods, 22. https://doi.org/10.1177/16094069231185456 |

| | |
|---|---|
| Reflective Writing | Meth P. ‘Coughing everything out’: the solicited diary method. In: *Qualitative Data : A Practical Guide To Textual, Media and Virtual Techniques*. ; 2017:94-116s. |
| Sorting | N/A |
| Space and Place Mapping | McGrath L, Mullarkey S, Reavey P. Building visual worlds: Using maps in qualitative psychological research on affect and emotion. Qualitative Research in Psychology. 2020 Jan 2;17(1):75-97. |
| Storytelling | Moreau KA, Eady K, Sikora L, Horsley T. Digital storytelling in health professions education: a systematic review. BMC medical education. 2018 Dec;18:1-9. |
| Temporal techniques | Sheridan J, Chamberlain K, Dupuis A. Timelining: visualizing experience. *Qual Res*. 2011;11(5):552-569. doi:10.1177/1468794111413235<br>Mignone J, Chase RM, Roger K. A graphic and tactile data elicitation tool for qualitative research : the life story board. *Forum Qual Sozialforsch*. 2019;20(2):1-26. |
| Vignette | Jenkins N, Bloor M, Fischer J, Berney L, Neale J. Putting it in context: the use of vignettes in qualitative interviewing. Qualitative research. 2010 Apr;10(2):175-98. |
| Walking | N/A |
